# Glutamine and NAA dissociate in ALS across somatotopically defined motor regions using 7T MRSI

**DOI:** 10.64898/2026.07.09.26357702

**Authors:** Zeinab Eftekhari, Sicong Tu, Timothy Ballard, Korbinian Eckstein, Bernhard Strasser, Fabian Niess, Lukas Hingerl, Wolfgang Bogner, Matthew C. Kiernan, Robert D. Henderson, Markus Barth, Thomas B Shaw

## Abstract

Amyotrophic lateral sclerosis (ALS) is increasingly understood as a progressive neurodegenerative disorder with distributed cortical and subcortical involvement, but *in vivo* metabolic mapping has been limited by the spatial coverage of single-voxel proton magnetic resonance spectroscopy (MRS). We acquired high-resolution whole-brain 7T 3D-CRT-FID-MRSI alongside motor-cortex single-voxel sLASER in five rapidly progressing people living with ALS (plALS) and seven non-neurodegenerative controls (NCs), with up to three sessions per participant. Regional metabolite ratios (N-Acetylaspartate [tNAA], glutamate [Glu], glutamine [Gln] to creatine [tCr], and Glu+Gln [Glx] to tNAA) were modelled with Bayesian hierarchical mixed-effects models, and the primary motor cortex was subdivided along its dorsoventral somatotopic axis (bulbar/face, hand/upper-limb, foot/lower-limb). At baseline, plALS showed a motor-cortex-selective tNAA/tCr deficit (motor composite −8.7%, 95% credible Interval [CrI] −16.1 to −1.1, posterior probability=0.99) accompanied by cortically diffuse glutamatergic elevation (Gln/tCr +25.6%, posterior probability=0.96; Glx/tNAA +10.4%, posterior probability=0.95). Reliable separation of the J-coupled glutamine and glutamate resonances at 7T revealed Gln/tCr as a more sensitive marker of glutamatergic dysregulation than Glu/tCr alone in this cohort. Within the somatotopic subdivision, all five plALS showed their peak Gln/tCr increase in the bulbar/face zone irrespective of clinical onset, including three lower-limb-onset patients. Annualised metabolite slope by zone correlated with the matched ALSFRS-R domain decline (Glx/tNAA r=0.82, p<0.001). Group-level longitudinal interactions were modest. Bayesian assurance simulations indicated Glx/tNAA as the most efficient candidate primary endpoint for a confirmatory cross-sectional study. These findings demonstrate that 7T whole-brain MRSI can resolve a metabolic dissociation between motor-selective neuronal compromised and somatotopically patterned glutamatergic dysregulation in ALS and provide design-ready endpoint and sample-size guidance for utility as a structural biomarker of brain function in clinical trials.

## 1. Introduction

Amyotrophic Lateral Sclerosis (ALS), the most common form of Motor Neuron Disease (MND), is a progressive neurodegenerative disorder marked by the degeneration of motor neurons in the brain and spinal cord, resulting in muscle weakness, atrophy, and respiratory failure. ALS affects both upper and lower motor neurons, leading to severe disability and reduced life expectancy.^1–4^ While the exact mechanisms underlying ALS remain unclear, metabolic dysregulation and excitotoxicity, driven by excessive glutamate activity, have been implicated in disease progression.^5^ These alterations, including disruptions in glutamatergic neurotransmission, neuronal integrity markers, and glial metabolism, highlight the need for advanced neuroimaging techniques to provide deeper insights into ALS pathophysiology.^6^

A particularly striking feature of ALS pathology is its somatotopic clinical presentation: symptoms typically begin focally in one body region (bulbar, upper limb, or lower limb) and progress regionally, often following the homuncular organisation of the primary motor cortex.^7,8^ Whether this clinical somatotopy is mirrored at the metabolic level and whether 7T MRSI can resolve such gradients within the motor strip, has not been systematically tested.

Proton Magnetic Resonance Spectroscopy (MRS)^9^ and Spectroscopic Imaging (MRSI)^10^ are non-invasive methods to explore neurochemical signatures in the brain.^11^ Both MRS and MRSI can detect metabolites relevant to ALS, including N-acetyl aspartate (NAA; a marker of neuronal health and function)^12^, choline-containing compounds (Cho; involved in membrane metabolism)^13^, creatine (Cr; related to cellular energy), myo-inositol (mIns; a marker of gliosis)^14^, glutamate (Glu; a primary excitatory neurotransmitter)^15^, and glutamine (Gln; a precursor to Glu and part of neurotransmitter cycling).^16^

Single Voxel Spectroscopy (SVS), most often implemented with MRI sequences such as semi-LASER (sLASER)^17,18^, acquires high-quality spectrum from a large (several cm^3^) predefined cubic volume of interest, making it well suited for targeted studies of specific structures, such as the motor cortex.^19–21^ However, it provides no spatial information beyond the selected voxel and may miss changes occurring elsewhere in the motor network. In contrast, MRSI samples multiple voxels across a larger field of view (FOV), enabling metabolic mapping of several cortical and subcortical regions simultaneously. This broader coverage can potentially capture the heterogeneous spread of ALS, where pathology often begins focally and asymmetrically and spreads to other regions.^22^ The trade-off is that MRSI typically has lower SNR per voxel and longer acquisition times, although modern acceleration and reconstruction methods have greatly improved its feasibility.^23–25^

The recent introduction of 3D-Concentric Ring Trajectory Free Induction Decay MRSI (3D-CRT-FID-MRSI) at ultra-high field (7T) enhances neurochemical detection due to its high signal-to-noise ratio (SNR) and spectral resolution.^26–29^ 7T MRSI offers clear advantages for studying ALS. The increased SNR improves sensitivity to small metabolic changes, while the improved spectral resolution aids in separating Glu and Gln, whose accurate quantification is important for probing excitotoxic mechanisms.^30–32^

Previous studies in plALS have consistently reported reductions in NAA and NAA ratios in motor cortex and subcortical regions^5,22,24,30,33,34,34–43^ and, in some cases, elevated Glu or Gln related to excitotoxicity,^30,32^ and some longitudinal studies that have reported inconsistent results,^39,42,44–54^ based on repeated spectroscopy measurements and within-subject changes over time or longitudinal slope in metabolite ratios. Exploratory MRSI studies in ALS have suggested abnormalities extending beyond the primary motor cortex, though these studies were performed mostly at 1.5T or 3T.^22,25,37,38,40,55–59^ 7T ALS studies have previously used either SVS or MRSI alone, rather than combining both within the same cohort, and with mixed outcomes.^24,34,58^ This study characterises both focal and network-level metabolic alterations over time in five plALS and seven age-matched non-neurodegenerative controls (NCs), and probes glutamatergic (Glu and Gln) differences in ALS over clinical dysfunction and onset locations. Additionally, we assess similarities in high-quality 7T SVS with high-resolution 7T 3D-CRT-FID-MRSI in plALS and include calculations for estimating the probability of meeting prespecified decision criteria in future studies to derive required sample sizes under clinically meaningful effect thresholds using MRSI.

All members of the plALS cohort have been deeply characterised (all rapidly progressing plALS, measured every 3 months) and recruited to confirm the utility of MRSI in plALS with clinical progression scales at the subject level. While this cohort is relatively clinically homogenous, at n=5, we here emphasise estimation over dichotomous significance testing, and report Bayesian probabilities of credible differences. We report baseline and longitudinal metabolite effects on clinically interpretable scales, including absolute differences in metabolite ratios and percent differences between groups (plALS vs NC) at baseline, as well as percent change over time, each with corresponding posterior uncertainty from a hierarchical Bayesian linear mixed-effects (LME) model. In addition, to facilitate subject-level interpretation and comparison across metabolites and regions, we provide a case-series approach to the spatial extent of metabolic alterations, mapped somatotopically to clinically relevant dysfunction.

Here, we also report subdivisions of the primary motor cortex along its dorsoventral somatotopic axis into bulbar/face, hand/upper-limb, and foot/lower-limb zones to test whether metabolic alterations track the homuncular organisation of clinical involvement, and translate the pilot effect estimates into Bayesian assurance curves to inform endpoint and sample-size selection for confirmatory MRSI studies.^60,61^

## 2. Material and Methods

### 2.1. Participants and study design

This study was conducted as a subset of the Biomarkers of Long surviving MND^62^ study at The University of Queensland, which enrolled participants across the spectrum of MNDs (for details, see ^62,63^). From the parent cohort, five people living with rapidly progressing classic ALS (plALS; mean age 56.9 +/− 7.9 years [range 46.5-66.2]; four male, one female) and seven age- and sex-matched non-neurodegenerative controls (NCs; mean age 55.9 +/− 7.0 years [range 47.6-67.7]; four male, three female) were included in the present analyses; group-level demographics and per-patient profiles are provided in Tables 1 and 2 (Section 3.1). plALS were recruited from the specialist MND neurology clinic at the Royal Brisbane and Women’s Hospital. All ALS diagnoses were confirmed by a consultant MND neurologist according to the Gold Coast diagnostic criteria for ALS,^64^ which provide a simplified, clinically applicable framework increasingly adopted in contemporary practice. King’s clinical stage was algorithmically derived from V1 ALSFRS-R subscale items per Balendra et al.^65^ All five plALS were Stage 2 at first scan.

**Table 1.** Group-level demographics and clinical profile at first scan. NC (n=7) includes one participant (sub-136) for whom the second visit served as the de-facto baseline owing to motion-related exclusion of the first visit. Continuous variables presented as mean ± SD [range]. King’s clinical stage was algorithmically derived from V1 ALSFRS-R subscale items.

| Variable | plALS ($n=5$) | NC ($n=7$) |
| --- | --- | --- |
| Sex (M:F) | 4:1 | 4:3 |
| Age at first scan (years), mean ± SD [range] | 56.9 ± 7.9 [46.5, 66.2] | 55.9 ± 7.0 [47.6, 67.7] |
| Site of onset (limb:bulbar) | 4:1 | n/a |
| Months from symptom onset to first scan, mean ± SD [range] | 27.2 ± 21.0 [11.2, 53.8] | n/a |
| Months from diagnosis to first scan, mean ± SD [range] | 16.4 ± 17.1 [2.2, 37.8] | n/a |
| Months from symptom onset to death, mean ± SD [range] | 46.7 ± 23.9 [28.3, 75.0] | n/a |
| ALSFRS-R total at first scan, mean ± SD [range] | 39.8 ± 4.4 [33, 44] | n/a |
| Forced vital capacity (% predicted), mean ± SD [range] | 86.6 ± 25.5 [67, 121] | n/a |
| TRICALS risk profile, mean ± SD [range] | -5.06 ± 0.40 [-5.4, -4.06] | n/a |
| King's clinical stage at first scan | Stage 2 in 5/5 | n/a |

**Table 2.** Individual clinical profiles for the five plALS at first scan, de-identified per ICMJE recommendations. King’s clinical stage was algorithmically derived from ALSFRS-R subscale items.

| ID | Sex | Age (banded years) | Onset site | Onset to scan (months) | ALSFRS-R | delta-FRS | King's | Visits | Follow-up (months) |
| --- | --- | --- | --- | --- | --- | --- | --- | --- | --- |
| ALS-1 | M | 50-54 | LL | 54 | 33 | 0.28 | 2 | 3 | 7 |
| ALS-2 | M | 60-64 | UL | 46 | 38 | 0.22 | 2 | 3 | 6 |
| ALS-3 | F | 65-69 | B | 12 | 44 | 0.34 | 2 | 3 | 8 |
| ALS-4 | M | 55-59 | LL | 11 | 41 | 0.62 | 2 | 2 | 3 |
| ALS-5 | M | 45-49 | LL | 13 | 43 | 0.38 | 2 | 3 | 7 |

Onset site: UL = Upper limb, LL = Lower limb, B = Bulbar.

To enrich the cohort for measurable disease progression within a feasible follow-up window, plALS were pre-screened on their TRICALS risk profile,^66^ a validated prognostic model integrating age, site of onset, diagnostic delay, ALSFRS-R slope, and respiratory function. Inclusion required a TRICALS score between −6 and 2, indicative of moderate-fast progression and a higher likelihood of detectable metabolic change over months rather than years. This deliberately narrowed the eligible pool: of seventeen ALS patient scans assessed against the TRICALS criterion and 7T compatibility (e.g., MRI safety, motion tolerance, no implanted dental ferromagnetic material), five plALS proceeded to the present spectroscopy sub-study, supporting a homogeneous fast-progressing phenotype amenable to longitudinal MRSI acquisition. Disease onset location was categorised as bulbar-onset, upper-limb-onset, or lower-limb-onset based on initial symptom presentation; three plALS had lower-limb-onset, one upper-limb-onset, and one bulbar-onset (Table 2).

At each visit, clinical data were collected for downstream analysis. Functional impairment was assessed using the ALS Functional Rating Scale-Revised (ALSFRS-R)^67^; respiratory function as the percentage of predicted forced vital capacity (%FVC)^68^. Disease-progression rate at first scan was calculated as (48 - ALSFRS-R) / months since symptom onset (delta-FRS)^69^.

Four plALS underwent three scanning sessions over 6 to 8 months and one underwent two scanning sessions over 3 months (rapid progression precluded a third session). Date of death was recorded for all five plALS. In the NC group, five of seven participants underwent two scanning sessions with an interval of 5 to 8 months; one NC completed only the first session (study withdrawal), and another NC’s first session was excluded for excessive head motion. All plALS were on Riluzole for at least 30 days prior to recruitment; no restrictions were placed on vital capacity at the time of inclusion. NC participants were screened with standard MRI safety procedures and excluded if they had a history of brain injury, neurosurgery, neurological disease, or psychiatric illness.

#### 2.1.1. Ethics statement

This study was approved by the local Human Research Ethics Committee (2021/HE000975). All participants provided written and informed consent to participate in research examining imaging biomarkers of MND and for deidentified data to be made available through publication and for other research purposes.

### 2.2. Image acquisition

This study was conducted using 7T imaging, a MAGNETOM 7T Plus MR scanner (Siemens Healthineers, Erlangen, Germany) with a 1-channel transmit/32-channel receive head coil (Nova Medical, Wilmington, MA, USA). All participants underwent an anatomical T_1_-weighted MP2RAGE scan for voxel positioning, cortical segmentation, atlas warping, and longitudinal alignment (Methods 2.3.2). The scanning parameters were as follows: at 7T, voxel size = 0.75×0.75×0.75 mm³, TR = 4300 ms, TE = 2.4 ms, TA = 6 mins, FA = 5°, matrix size = 192×224×256.^70,71^

A high-resolution whole brain 3D-CRT-FID-MRSI was acquired with the following parameters^29^: voxel size = 3.4×3.4×3.5 mm³, TR = 460 ms, TE = 1.3 ms, TA = 14:30 mins, FA = 39° (calculated as the nominal average Ernst angle for NAA, tCr, tCho, Glu, and mIns), matrix size = 64×64×31, FOV = 220×220×110 mm³, Volume of Interest [VOI] = 220×220×60 mm³, readout duration = 311 ms, spectral bandwidth = 2778 Hz. 3D-CRT-FID-MRSI has an ultrashort acquisition delay (∼1.3 ms), which minimizes T_2_ and J-evolution-related SNR loss; while eliminating refocusing pulses reduces specific absorption rate (SAR).^26,72–75^ No non-suppressed water spectra were acquired due to time constraints, though interleaved water-calibration were acquired.^76^ The protocol also included B1+ mapping for flip-angle optimisation and variable temporal interleaves (1-3, depending on ring radii)^77^ to maintain spectral bandwidth for larger readout circles and reduce scan times further.^29^ An imaging slab with a thickness of 60 mm was placed parallel to the horns of the corpus callosum for MRSI acquisitions. No lipid suppression was applied to allow for a shorter TR. Full sequence details are available in.^19^

All SVS data were acquired after B_0_ shimming via FAST(EST)MAP,^75,78^ optimisation of RF transmitter power, and calibration of VAPOR water suppression.^79^ An sLASER-SVS sequence with the following parameters was used^17,18^: voxel size = 25×25×25 mm³, TR = 8000 ms, TE = 26 ms, 16 averages, TA = 2.5 mins. Full sequence details are described in our previous work.^20^ For each participant the voxel was consistently placed in the right precentral gyrus (representing the area corresponding to the upper limb region of the motor homunculus (See Figure 1).^80^

**Figure 1.**
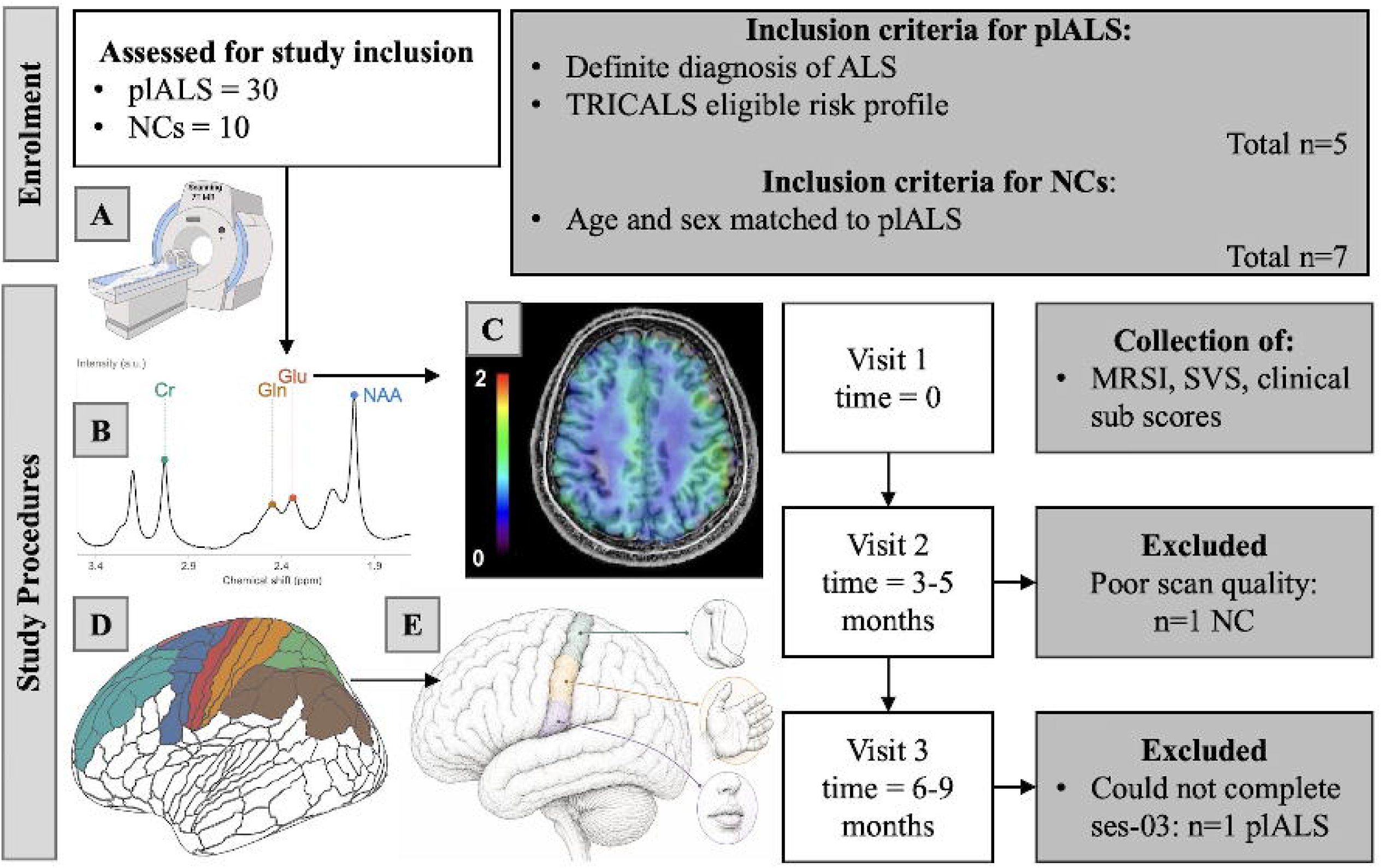
Overview of study design and data acquisition workflow. Participants originally from the larger cohort of 30 plALS and 10 NCs were chosen for this sub-study. Five people living with amyotrophic lateral sclerosis (plALS) and seven age- and sex-matched healthy controls were included. plALS were classified as eligible for inclusion based on their TRICALS risk profile scores. Each participant underwent 2-3 sessions on a 7T MRI scanner (A). Clinical data were collected at each visit. Metabolite data were collected (B) using large cortical coverage MRSI (C), enabling region-wise metabolite mapping (D), while SVS was acquired in the precentral gyrus. Spectra were quantified for metabolites including total N-acetylaspartate (tNAA), creatine (Cr), glutamate (Glu), and glutamine (Gln), and the combination of Gln and Glu known as Glx (B). Somatotopic regions (E) were defined by splitting motor regions into thirds for bulbar, arm, and leg region-specific MRSI profiles.

### 2.3. Data Post-processing and Metabolite Quantification

The MRSinMRS checklist can be found in Table S1 in supplementary material.^81^

#### 2.3.1. Spectroscopic Data

MRSI data were processed offline using a custom-developed pipeline and all sLASER-SVS post-processing was performed in Osprey (v.2.5.0), an open-source MRS analysis toolbox.^,82^ The automated pipeline included standard steps for spectral pre-processing, quality control, and quantification. Detailed descriptions of the full post-processing workflow are provided in our previous works.^19^ Please find detailed methodology regarding spectral quality evaluation in Supplementary Material section 7.3.

Metabolite concentrations were quantified using LCModel, with spectral fitting applied to different chemical shift ranges depending on the acquisition method.^84^ The range for MRSI was restricted to 1.8 to 4.2 ppm to avoid lipid contamination at 1.3 ppm. The LCModel basis sets for both SVS and MRSI included 19 simulated metabolites and a measured macromolecule spectrum as previously reported.^83,85–87^ Since absolute metabolite concentrations could not be determined due to the lack of localized T_1_ relaxation times and an internal water reference, all metabolite values were normalized to tCr. In addition, the combination of Glu and Gln (Glx) was normalized to total N-acetylaspartate (tNAA), in accordance with previous MRSI studies in ALS. This complementary referencing was used because reductions in tNAA may reflect neuronal loss, and normalising excitatory metabolites to tNAA can help distinguish changes related to excitotoxic processes from those driven by volume loss.^30^

#### 2.3.2. Anatomical Segmentation and ROIs

T1-weighted MP2RAGE images were processed using a longitudinal FastSurfer pipeline.^88^ For each participant with multiple sessions, an unbiased subject-specific template was constructed across all available timepoints, and white-matter and grey-matter segmentations were performed jointly across sessions to maximise within-subject consistency over time. The resulting subject-specific brain models then served as a common space into which the Glasser multi-modal cortical parcellation(180 Regions of Interest [ROIs] per hemisphere)^89^ was warped using a sequence of linear and nonlinear registrations,^90,91^ yielding longitudinally consistent Glasser ROI labels in each participant’s native T1-weighted space at every timepoint.

Prior to referencing each metabolite to total creatine (tCr) or to tNAA, for the Glx/tNAA ratio, each individual metabolite map was smoothed using a biharmonic spatial smoother to suppress voxel-level noise while preserving regional spatial structure.^92^ Segmentations were then resampled to the MRSI map grids and the four metabolite ratios (tNAA/tCr, Glu/tCr, Gln/tCr, Glx/tNAA) were computed per voxel.

ROI-level extraction proceeded by intersecting the atlas parcels with the MRSI volume of interest (VOI) and the FastSurfer grey-matter mask. To minimise partial-volume effects, only voxels lying within both the cortical grey-matter mask and the VOI were retained, and voxels with invalid metabolite estimates (Cramer-Rao Lower Bound (CRLB) > 50%)^93^ were excluded. We did not apply explicit tissue-fraction correction (for example, regression on Grey Matter (GM), White Matter (WM), and Cerebrospinal Fluid (CSF) fractions or CSF-fraction scaling), so metabolite-ratio estimates within each ROI are interpreted as reflecting the dominant grey-matter composition of that anatomically defined region. White-matter regions were excluded from the present analyses to keep the focus on cortical somatotopic mapping. Per-voxel Interquartile Range (IQR) outlier filtering (k = 1.5, pooled across groups within each ROI)^94^ was applied to remove residual artefactual extremes prior to modelling.

For the principal cross-sectional analyses, a ten-ROI set was retained: four motor-network ROIs (Primary Motor [parcels 4 and 3a], Supplementary Motor Area[SMA; 6ma, 6mp, SCEF], Premotor [6a-v, 55b, FEF, PEF], Cingulate Motor [23c, 24dd, 24dv]) and six non-motor comparator ROIs (Superior Parietal [7AL, 7Am, 7PC, 7PL, 7Pl, 7Pm, AIP, LIPd, LIPv, MIP, VIP], Somatosensory [1, 2, 3b, 5L, 5m, 5mv], Dorsolateral Prefrontal, Anterior Cingulate / Medial Prefrontal, Inferior Parietal, Posterior Cingulate). A pre-specified motor composite endpoint was derived by pooling posterior percent-difference draws across the four core motor ROIs (Primary Motor, SMA, Premotor, Cingulate Motor); Somatosensory and Superior Parietal were retained as motor-adjacent regions and reported separately. For somatotopic analyses (3.6), the Primary Motor parcel set was further subdivided into superior, middle, and inferior thirds along the dorso-ventral axis to approximate the bulbar/face, hand/upper-limb, and foot/lower-limb regions of the homunculus.

### 2.4. Statistical Analysis

All models were fitted in a Bayesian hierarchical framework using brms with the cmdstanr backend, implemented in R on the voxel-level data.^95–98^ Working directly on individual retained voxels rather than collapsing to a single ROI mean per participant allows the within-ROI spatial variability — the very signal that high-resolution MRSI is acquired to capture — to inform the posterior, an important gain in a small cohort. All models used a Student-t likelihood rather than the conventional Gaussian so that occasional outlier voxels (residual lipid contamination, line-broadening, local susceptibility) contributed appropriately less leverage to the posterior; the degrees-of-freedom parameter was estimated from the data. Given the small cohort size and the nested structure of voxel-level MRSI data, all models were estimated within a Bayesian hierarchical framework. This allowed us to account for variability between participants and across ROIs, while also examining the communalities across them (via the fixed effects).

#### Cross-sectional models

For each metabolite ratio we fitted a bilateral baseline model (value ∼ Group + (1 | Participant) + (1 + Group || ROI)) over the ten-ROI set, with a participant-level random intercept and ROI-varying random intercept and group slope. A separate lateralised baseline model added hemisphere as a fixed effect (value ∼ Group * Hemisphere + (1 | Participant) + (1 + Group * Hemisphere || ROI-at-baseline)) restricted to a 6-ROI subset (5 motor regions plus Dorsolateral Prefrontal as a non-motor anchor) to preserve estimation stability when doubling the ROI dimensionality with hemisphere.

#### Longitudinal model

For each metabolite ratio we fitted value ∼ Timepoint * Group + (1 + Timepoint | Participant) + (1 + Timepoint + Timepoint:Group || ROI), where Timepoint is months since first scan (mean-centred). The fixed Group:Timepoint interaction estimated the population-average difference in rate of change between plALS and NCs; ROI-varying interaction terms allowed regional heterogeneity in slope differences.

Reporting Glx alongside separately quantified Glu and Gln took advantage of the lower CRLB for the combined Glx signal compared with Gln aloneand preserved comparability with prior 1.5T and 3T ALS MRS work.^,30,43^

#### Posterior summarisation

Group differences were computed directly from the posterior draws to preserve uncertainty. For each posterior sample, we computed the implied NC mean and the implied plALS-minus-NC difference for each ROI. This difference was expressed as a percentage of the NC mean and only then summarised using the posterior median and 95% credible interval. Pre-specified motor composite endpoints, the relevant ROI-specific draws, were averaged first before posterior distribution summarisation. Full prior specifications, sampling parameters, and convergence diagnostics are provided in Supplementary Methods S1.

#### Bayesian assurance

To support trial design, we performed simulation-based Bayesian assurance analyses for the motor-composite endpoints, per metabolite ratio.^60^ Posterior draws from the pilot motor-composite model were used to generate plausible “true” effect sizes; baseline datasets were simulated at candidate sample sizes (10 to 60 per group); a model was refit to each simulated dataset; and a study was deemed successful if the posterior probability that the percent difference exceeded each of three thresholds (delta = 2%, 5%, 10%) was at least 0.95 under a one-sided rule aligned with the sign of the pilot effect. Assurance was defined as the proportion of successful simulations at each candidate sample size. Simulation details are in Supplementary Methods S1.4.

### 2.5. Comparison of MRSI and MRS (single voxel) trends in plALS

Previous work from our group has demonstrated that both MRSI and single voxel sLASER acquisitions provide reliable and comparable metabolite estimates across a range of regions and acquisition/processing choices.^19^ Here, we performed a qualitative cross-method comparison between sLASER-SVS acquired in the motor cortex and MRSI-derived estimates from the anatomically corresponding precentral gyrus ROI.

Because sLASER-SVS and MRSI differ substantially in sampling (single voxel versus many voxels), spatial coverage, and acquisition/processing parameters, we did not treat these data as directly interchangeable or attempt formal equivalence testing. Instead, for each metabolite ratio of interest, we summarised group-level differences for each modality using z-scored contrasts (plALS vs NC) within the motor cortex/precentral region. This visualisation emphasises relative direction and magnitude of group effects (increases versus decreases) across modalities while avoiding over-interpretation of absolute agreement.

## 3. Results

### 3.1. Participant Demographics and Clinical Profiles

Five plALS and seven NCs were included in the analyses (Table 1). Groups were comparable in age and sex distribution. All five plALS were classified as fast-progressing and well-characterised at recruitment (TRICALS risk profile −5.8 to −4.7)^66^, reflecting an a priori selection criterion intended to maximise the likelihood of detecting metabolic change within the available follow-up window (see Methods 2.1). All five plALS met the Gold Coast Criteria for the diagnosis of ALS.^64^ All five plALS subsequently passed away (median time from first scan to death 17.0 months, range 14.0 to 24.7).

Given the small ALS sample, individual clinical profiles are summarised in Table 2 (de-identified by age-band reporting and omission of side-of-onset and predominance). The cohort included three lower-limb-onset, one upper-limb-onset, and one bulbar-onset case. Baseline ALSFRS-R ranged 33 to 44 (mean 39.8, SD 4.4) and the disease-progression rate (delta-FRS) ranged 0.22 to 0.62 per month (mean 0.37, SD 0.15), spanning the slow-to-fast spectrum within the TRICALS-eligible cohort. King’s clinical stage, derived algorithmically from the ALSFRS-R subscale items,^65^ was Stage 2 in all five plALS at first scan (regional involvement of two of three motor regions: bulbar, upper limb, or lower limb, without nutritional or respiratory failure).

### 3.2. Data quality and MRSI validation

Spectral quality was comparable between plALS and NCs at both the voxel and ROI level, with detailed SNR, Full-Width Half-Maximum (FWHM), and CRLB summaries provided in Supplementary Materials (see Supplementary Results S2.1). Building on prior validation of this MRSI pipeline against SVS in healthy controls,^19^ to support the validity of MRSI-derived metabolite ratios in this cohort, we compared MRSI estimates against SVS acquired in the same imaging session, restricting the comparison to a representative ROI (right primary motor cortex) and to median values per participant per session.

Although MRSI estimates are inherently more variable owing to the larger number of voxel-level measurements per participant, the two acquisitions showed substantial overlap to SVS within each cohort across all four metabolite ratios of interest (Figure 2). The expected direction of effect in plALS was concordant between methods: tNAA/tCr and Glu/tCr were lower, while Gln/tCr and Glx/tNAA were higher in plALS relative to NCs in both MRSI and SVS measurements. Together these findings support the use of MRSI-derived metabolite ratios for the regional analyses that follow.

**Figure 2.**
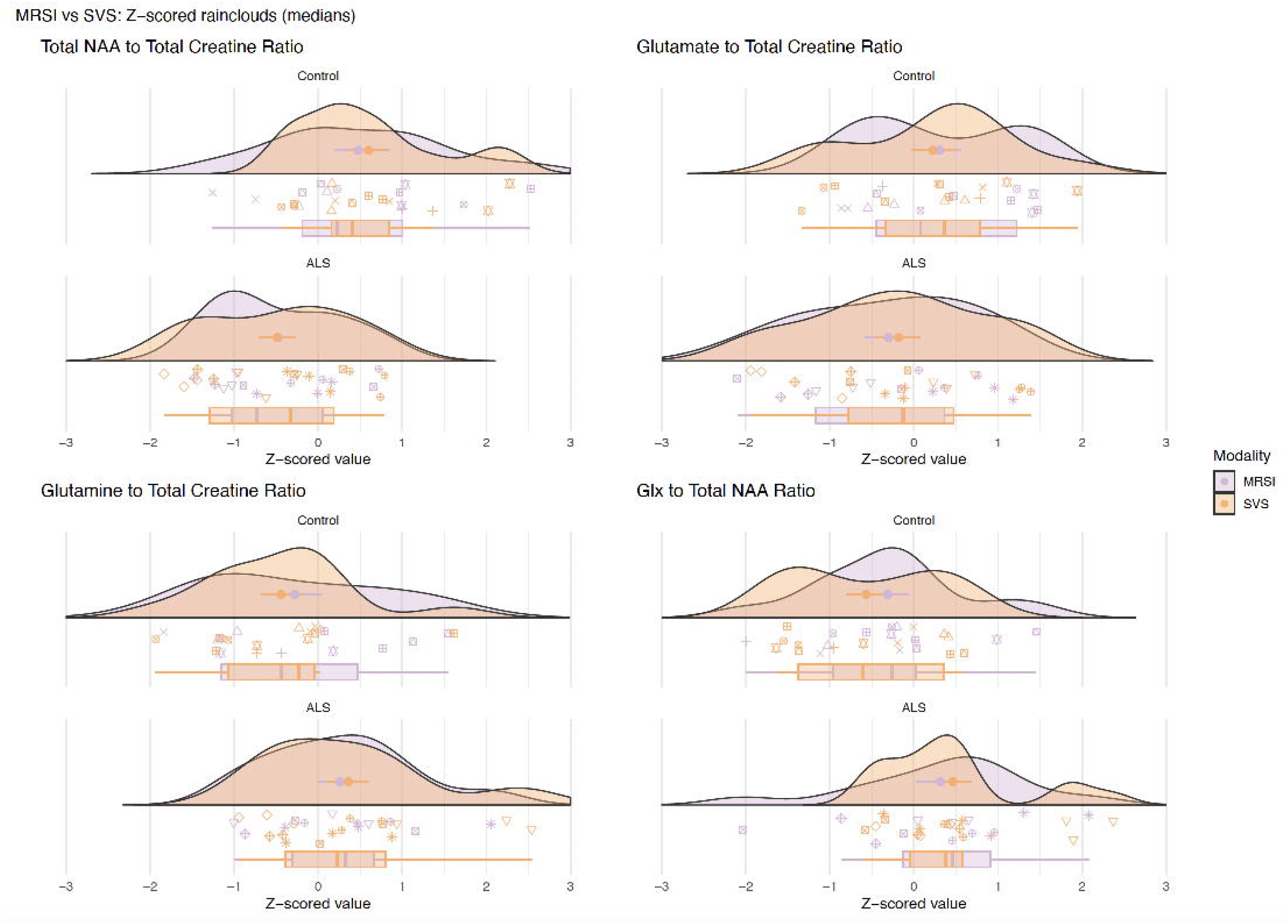
Raincloud plots of Z-scored metabolite ratios (tNAA/tCr, Glu/tCr, Gln/tCr, Glx/tNAA) from MRSI and SVS in the right primary motor cortex. MRSI values represent multiple voxel measurements per participant summarised as medians; SVS provides a single voxel measurement per participant per session, hence MRSI shows greater within-subject spread. All four ratios show concordant directionality between modalities, with tNAA/tCr and Glu/tCr reduced and Gln/tCr and Glx/tNAA elevated in plALS relative to NCs.

### 3.3. Cross-sectional metabolite differences across the motor network

Bayesian hierarchical mixed-effects models (Methods 2.4)^96^ estimated baseline group differences in metabolite ratios across a ten-ROI cortical set: six motor-network ROIs (Primary Motor [areas 4, 3a], Somatosensory [1, 2, 3b, 5L/m/mv], SMA [6ma, 6mp, SCEF], Premotor [6a-v, 55b, FEF, PEF], Cingulate Motor [23c, 24dd, 24dv], Superior Parietal) and four non-motor comparator ROIs (Dorsolateral Prefrontal, Anterior Cingulate / Medial Prefrontal, Inferior Parietal, Posterior Cingulate). A pre-specified motor composite endpoint was derived by pooling posterior percent-difference draws across the four core motor ROIs (Primary Motor, SMA, Premotor, Cingulate Motor); Somatosensory and Superior Parietal are reported separately as motor-adjacent regions. Effects are reported as posterior median percent differences (plALS vs NCs) with 95% credible intervals (CrI) and the posterior probability (denoted as p(0|X) that the effect lay in the indicated direction (Figure 3). At the motor-composite level, tNAA/tCr was reduced by 8.7% in plALS (95% CrI −16.1 to −1.1, p(0|X) (<0) = 0.99), Gln/tCr was elevated by 25.6% (95% CrI −4.4 to +61.5, p(0|X) (>0) = 0.96), Glx/tNAA was elevated by 10.4% (95% CrI −2.5 to +25.6, p(0|X) (>0) = 0.95), and Glu/tCr was directionally reduced by 4.5% with weaker evidence (95% CrI −17.9 to +11.5, p(0|X) (<0) = 0.75). See Figure 3 and Table 3 for full details, including motor region breakdowns, and Supplementary Table S5 for non-motor regions.

**Figure 3.**
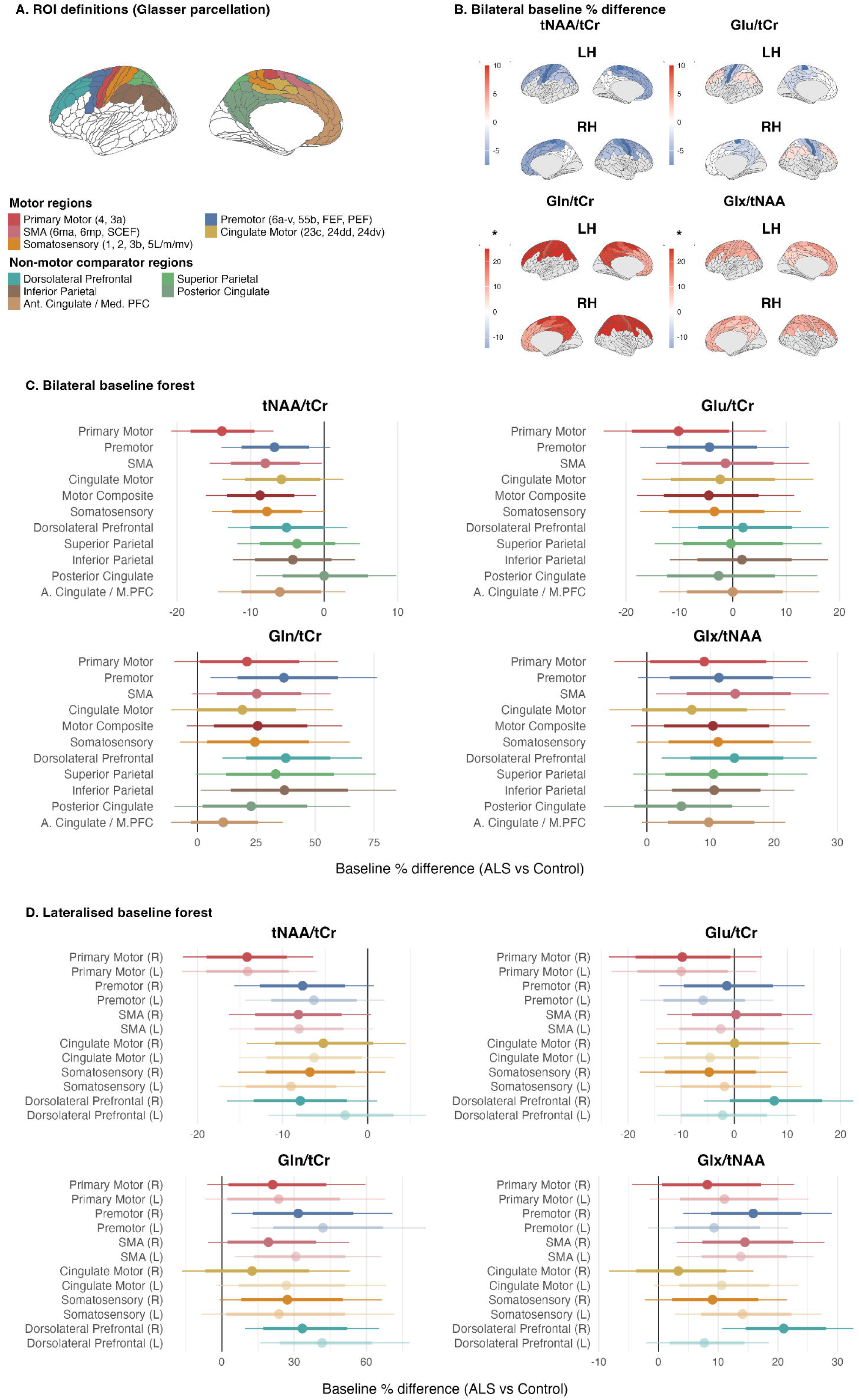
Baseline metabolite differences across the cortex. **(A)** ROI definitions based on the Glasser parcellation: motor-network ROIs (Primary Motor, Somatosensory, SMA, Premotor, Cingulate Motor, Superior Parietal) and non-motor comparator ROIs (Dorsolateral Prefrontal, Anterior Cingulate / Medial Prefrontal, Inferior Parietal, Posterior Cingulate) Colours match effects in panels C and D. **(B)** Bilateral baseline percent difference (plALS minus NCs) across the four metabolite ratios, shown on the Glasser cortex with a diverging colour scale* (blue indicates lower in plALS, red indicates higher in plALS). **(C)** Bilateral baseline forest plot showing posterior median and 95% credible interval for all four metabolite ratios across the ROI set, with motor composite endpoints highlighted. **(D)** Lateralised baseline forest plot showing the same metabolite set with left and right hemispheres separated. Colours are matched across hemisphere, use panel A as a guide.

**Table 3.**
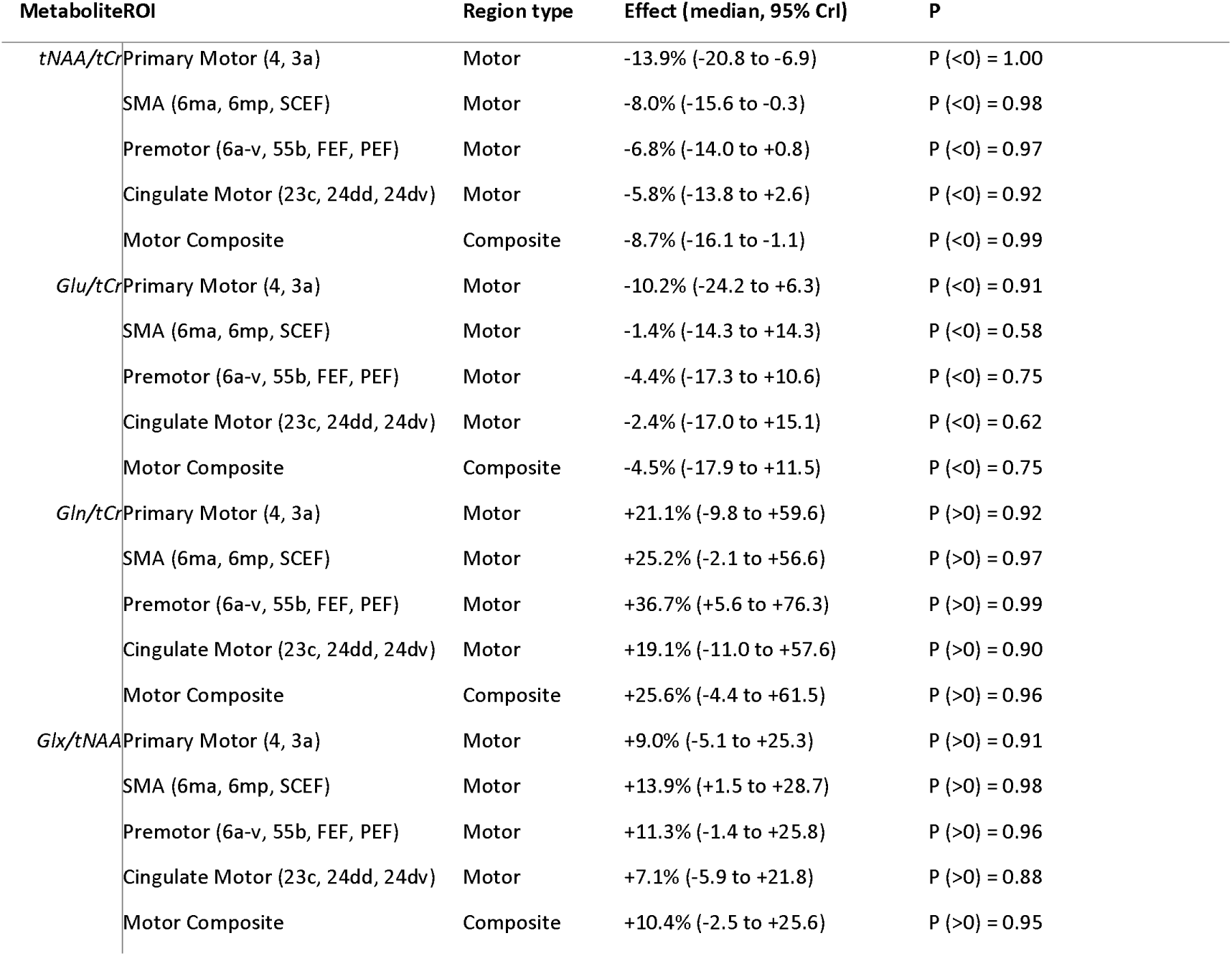
Baseline ROI-wise group differences (plALS minus NCs, expressed as percent of NC mean) from Bayesian hierarchical mixed-effects models with Student-t likelihood. Motor composite estimates are derived by averaging ROI-specific posterior percent-difference draws across the six motor ROIs. P denotes the posterior probability of group separation in the indicated direction.

The tNAA/tCr deficit was anatomically focal: it was largest in Primary Motor cortex (−13.9%, 95% CrI −20.8 to −6.9), preserved across all four ROIs comprising the motor composite, and substantially attenuated in the four non-motor comparators (mean median difference −8.6% across core motor ROIs vs −4.5% across non-motor comparators; Posterior Cingulate effectively unchanged at 0.0%, 95% CrI −9.2 to +9.8). This pattern is consistent with motor-cortical neuronal compromise dominating the tNAA/tCr signal in plALS at this disease stage. The directionally reduced Glu/tCr was strongest at Primary Motor (−10.2%, 95% CrI −24.2 to +6.3) but credible intervals included zero across all ROIs.

By contrast, the elevations in Gln/tCr and Glx/tNAA were not motor-selective. Gln/tCr was elevated to a similar degree in motor and non-motor cortex (motor mean +25.5% vs non-motor +27.7%) and Glx/tNAA likewise (+10.3% vs +10.2%), with the Dorsolateral Prefrontal cortex showing the largest Gln/tCr increase of any ROI (+37.5%, 95% CrI +10.7 to +69.9), see Table 3 and Figure 3. This dissociation, motor-selective tNAA/tCr loss alongside cortically diffuse glutamatergic elevation, suggests that glutamatergic dysregulation in this cohort extends beyond the regions of greatest neuronal compromise. Notably, the posterior probability of group separation was higher for Gln/tCr (p(0|X) = 0.96) than for Glu/tCr (p(0|X) =0.75), consistent with 7T FID-MRSI providing reliable separation of the J-coupled Gln and Glu peaks and revealing Gln/tCr as a more sensitive marker of glutamatergic dysregulation than Glu/tCr alone in this cohort.

### 3.4. Hemispheric lateralisation

To probe whether the cross-sectional metabolite differences were asymmetric across hemispheres, we fitted a separate lateralised baseline model that added hemisphere as a fixed effect and a Group x Hemisphere interaction at the ROI level. To preserve estimation stability at n=12 participants and avoid doubling the random-effect dimensionality across all ten cortical ROIs, this lateralised model was restricted to the five motor ROIs (Primary Motor, Somatosensory, SMA, Premotor, Cingulate Motor) and a single non-motor anchor (Dorsolateral Prefrontal); the bilateral model reported in 3.3 retains the broader ten-ROI set for comprehensive cortical coverage (Figure 3, panels B-D).

The tNAA/tCr deficit was strikingly symmetric in the Primary Motor cortex (right-14.2%, left −14.1%) and SMA (right −8.1%, left −8.1%), with only modest asymmetry in the non-motor anchor (right −7.9%, left −2.7%; mean across all six ROIs in this model: right −8.3%, left −7.7%). Glu/tCr and Glx/tNAA were likewise approximately symmetric at the Primary Motor cortex (Glu/tCr right −9.8%, left −10.0%; Glx/tNAA right +8.2%, left+11.0%) and across the broader ROI set in this model (Glx/tNAA mean right +12.0%, left+11.1%). Gln/tCr showed a consistent but modest left-greater pattern across multiple ROIs (mean right +24.2%, left +31.4%; difference 7.2 percentage points), most pronounced in Cingulate Motor (−14.2 pp), SMA (−11.5 pp) and Premotor (−10.2 pp) cortex. With only five plALS, of whom two had right-side limb onset, two left-side, and one bulbar, this dataset cannot resolve whether the Gln/tCr asymmetry reflects a true neurobiological pattern or sampling variation, and we treat it as a hypothesis-generating observation pending replication in a larger cohort.

### 3.5. Longitudinal group-level trajectories

For longitudinal modelling, voxel-level Bayesian hierarchical models added a Group x Time fixed-effect interaction with participant-level random intercepts and slopes and ROI-varying intercepts, time slopes, and Group x Time interactions (Methods 2.4). At the population-average level, fixed-effect Group x Time interactions were small and uncertain for all four metabolites (tNAA/tCr 0.01 [95% CrI −0.02, +0.04]; Glu/tCr 0.00 [−0.02, +0.02]; Gln/tCr −0.01 [−0.02, +0.01]; Glx/tNAA −0.01 [−0.03, +0.02]). The motor-composite annualised interaction slopes were likewise indecisive (tNAA/tCr +0.19 [−0.17, +0.56]; Gln/tCr −0.07 [−0.22, +0.08]; Glx/tNAA −0.12 [−0.40, +0.15]; Glu/tCr +0.01 [−0.25, +0.28]). The strongest ROI-specific signals were not in the motor cortex but in non-motor regions, with Gln/tCr declining faster in plALS than NCs in Superior Parietal (−0.16/yr [−0.31, −0.01], p(0|X) (<0)=0.98) and Dorsolateral Prefrontal (−0.14/yr [−0.29, +0.01], p(0|X) (<0)=0.97), and Glx/tNAA declining faster in Anterior Cingulate / Medial Prefrontal (−0.27/yr [−0.55, 0.00], p(0|X) (<0)=0.97) and Dorsolateral Prefrontal (−0.24/yr [−0.51, +0.04], p(0|X) (<0)=0.96). Whether these declines reflect genuine changes in glutamatergic concentrations or denominator drift in tCr cannot be resolved at the group level in this dataset, as a group-level longitudinal tCr model was not fitted; per-zone tCr stability for the five plALS is examined directly in the case series (3.6) and in Supplementary Materials.

With five plALS followed for a median of 7 months, the present cohort is underpowered to resolve consistent rate-of-change differences at the population level; full Group x Time interactions and ROI-level slopes for all four metabolites are reported in Supplementary Table S2 and Supplementary Figure S1. Individual patient trajectories (3.6) provide a more informative view of within-cohort heterogeneity over the same follow-up.

### 3.6. Somatotopic metabolite-function mapping

To explore whether the regional metabolic changes reflect ALS pathology in a clinically interpretable way, we split the motor cortex into superior, middle, and inferior thirds and pooled voxels within each third into bulbar/face, hand/upper-limb, and foot/lower-limb zones, mirroring the homuncular organisation of the primary motor cortex.^7,8^ Within each plALS, we extracted the annualised metabolite slope per zone across the available scans and compared it against the corresponding ALSFRS-R domain subscore trajectory (bulbar items 1-3, upper-limb items 4-6, lower-limb items 8-9)^67^, yielding 15 patient-zone observations across the cohort.

The most striking pattern was for Gln/tCr. Across the cohort, the median annualised Gln/tCr change differed monotonically across the somatotopic axis: +18.2% in the bulbar/face zone, −19.2% in the hand zone, and −32.6% in the foot zone (Figure 4B). All five plALS showed their peak Gln/tCr increase in the bulbar/face zone irrespective of clinical onset site (Figure 4A; the same bulbar-predominant pattern was visible for Glx/tNAA, while tNAA/tCr and Glu/tCr showed more variable per-patient patterns), including the three lower-limb-onset patients (ALS-1 +18.2% peak bulbar; ALS-4 +8.4%; ALS-5 +18.2%; bulbar-onset ALS-3 +27.4%; upper-limb-onset ALS-2 +30.7%). This bulbar-predominant Gln/tCr elevation occurred even in patients whose clinical decline was dominated by limb dysfunction, suggesting a topographic gradient in glutamatergic metabolism that is not strictly tied to the initial site of clinical involvement.

**Figure 4.**
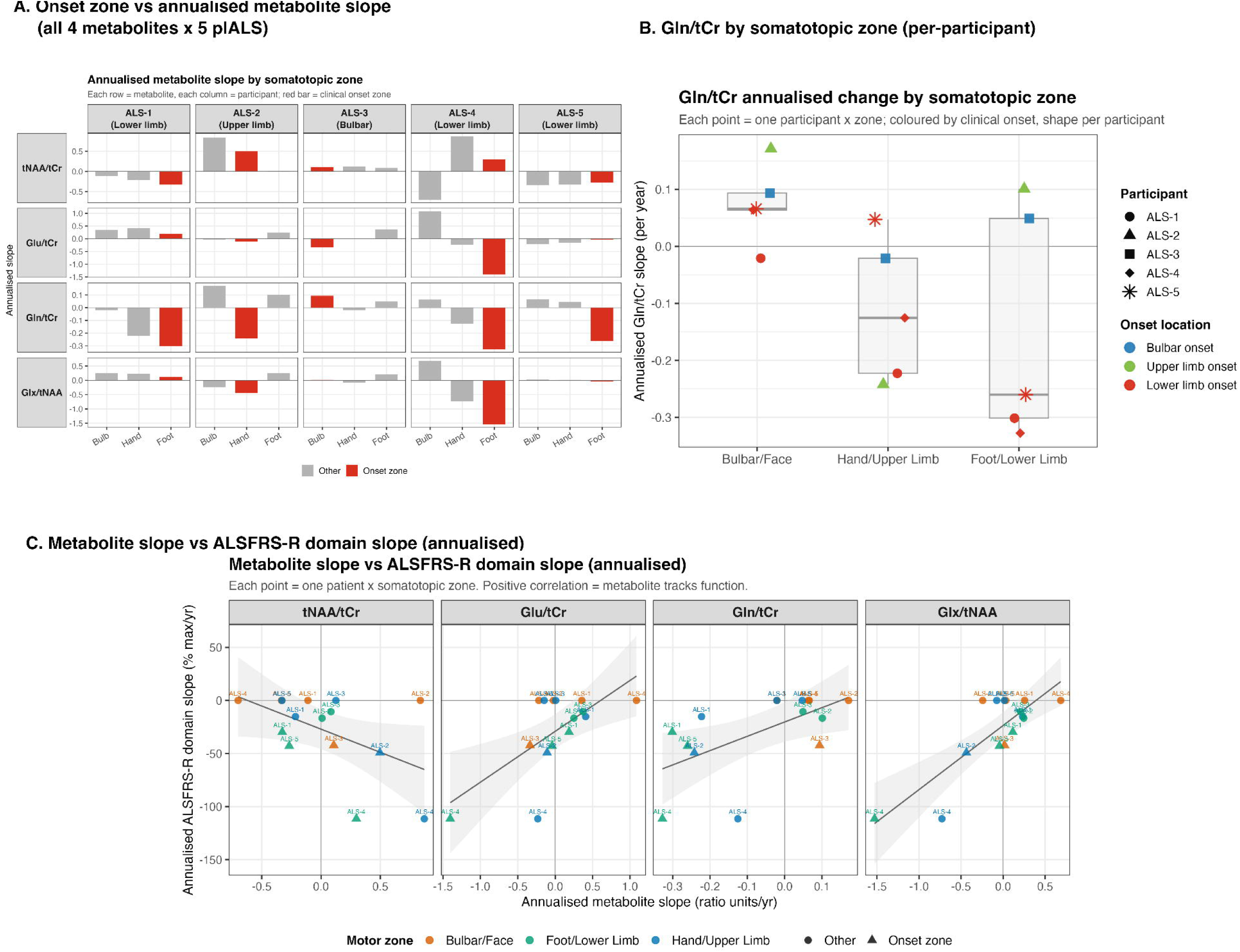
Somatotopic metabolite-function mapping in the five plALS. **(A)** Annualised metabolite slope by somatotopic zone for each plALS, shown as a 4-metabolite (rows) by 5-participant (columns) grid; the bar corresponding to each patient’s clinical onset zone is highlighted in red. Across patients, Gln/tCr peaks consistently in the bulbar/face zone irrespective of clinical onset, and the same is broadly true for Glx/tNAA; the tNAA/tCr and Glu/tCr panels show more variable patterns across patients. **(B)** Gln/tCr annualised change by somatotopic zone, with one point per participant per zone; points are coloured by clinical onset (bulbar / upper-limb / lower-limb) and shaped by participant (ALS-1 to ALS-5). **(C)** Per-zone metabolite slope plotted against the matched ALSFRS-R domain sub score slope across the five metabolite ratios examined; Glx/tNAA shows the strongest coupling (r=0.82, p<0.001), with positive slope corresponding to slower functional decline in the matched body region.

Within the same 15 patient-zone observations, annualised metabolite slopes correlated with the corresponding ALSFRS-R domain subscore slope as follows: Glx/tNAA r=0.82 (p<0.001), Glu/tCr r=0.67 (p=0.006), Gln/tCr r=0.60 (p=0.019), tNAA/tCr r=-0.51 (p=0.052), and tCr r=-0.73 (p=0.002). Glx/tNAA was the strongest metabolite-function coupling, with greater (less negative) slope corresponding to slower functional decline in the matched body region (Figure 4C). We note that these correlations treat each of the 15 patient-zone pairs as independent, whereas they are clustered within five patients; the same direction and rank ordering held in a sensitivity analysis using participant-mean slopes (Supplementary Table S4).

Because all metabolite ratios are referenced to tCr (or to tNAA in the case of Glx/tNAA), we examined whether tCr itself drove the somatotopic patterns. Median annualised tCr change by zone was +2.0% (bulbar/face), +7.5% (hand), and +11.2% (foot), a gradient of approximately 9 percentage points across zones, opposite in direction and substantially smaller than the ∼50 percentage-point Gln/tCr gradient (per-patient tCr trajectories shown in Supplementary Figure S2). The tCr-function correlation (r=-0.73) reflects this denominator drift, and absolute metabolite quantification will be important in future work; nonetheless, the magnitude of the Gln/tCr gradient is unlikely to be explained by tCr alone, and the bulbar-predominant Gln peak persists when each patient’s tCr trajectory is accounted for.

Per-patient longitudinal trajectories of motor-composite metabolites and ALSFRS-R sub scores are detailed in Supplementary Figures S1-S3 and Supplementary Table S3; the cohort exhibits substantial inter-individual heterogeneity in metabolite trajectories over the 3-8-month follow-up window, motivating individual rather than population-average endpoints in this rare-disease context. Per-participant, per-session voxel-level distributions for all four metabolite ratios, alongside matched ALSFRS-R domain trajectories and clinical progression metrics, are shown in Supplementary Figure S3.

### 3.7. Bayesian assurance for prospective trial design

To inform the design of a follow-up cross-sectional study using the same 7T 3D CRT FID-MRSI pipeline, we performed Bayesian assurance analyses for the four metabolite-composite endpoints (Methods 2.4.2).^60,61^ Posterior draws from the pilot motor-composite model were propagated through parametric simulation across candidate sample sizes (10 to 60 per group); see Methods 2.4 and Supplementary Methods S1.4 for the full simulation procedure (Figure 5).

**Figure 5.**
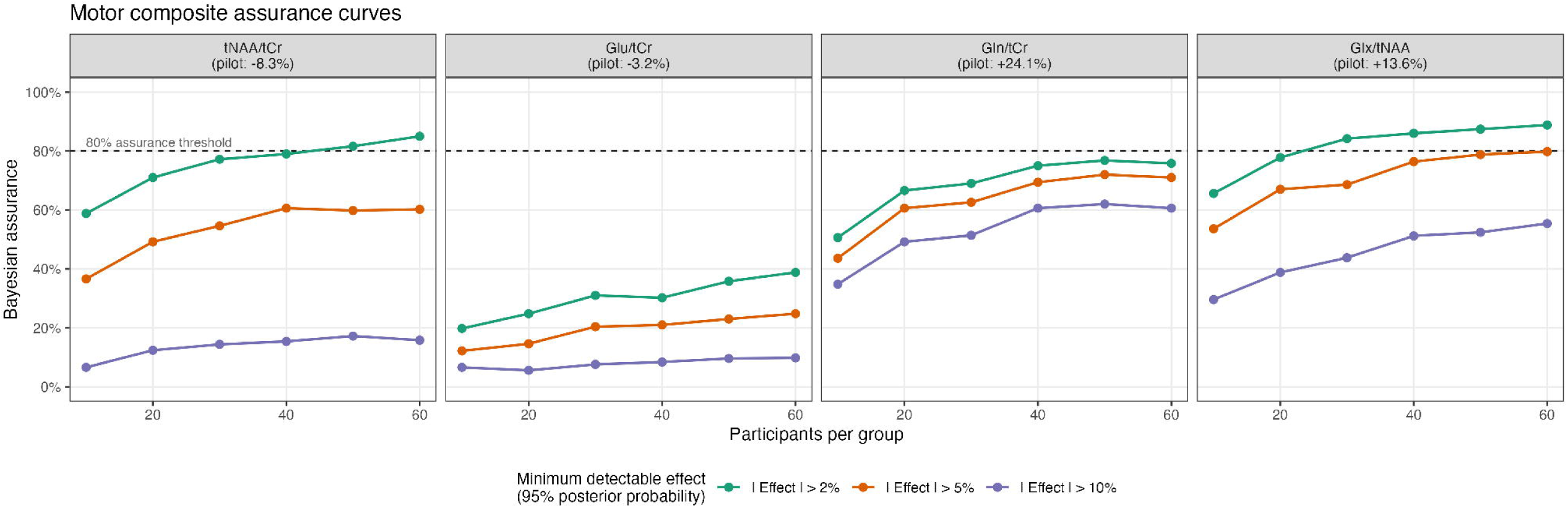
Bayesian assurance curves for the metabolite motor-composite endpoints. For each metabolite, the y-axis shows the simulation-estimated probability of declaring a successful study (posterior probability of group difference exceeding delta at least 0.95) as a function of per-group sample size. Three thresholds are shown: |delta| > 2% (clinically modest), > 5% (clinically meaningful), and > 10% (large). Direction of the test is set by the sign of the pilot motor-composite effect. Glx/tNAA approaches 80% assurance at N approximately 30 per group for the 2% threshold; tNAA/tCr at approximately 50; Gln/tCr approaches but does not exceed 80% within the simulated range (maximum 76% at N=60).

Among the four endpoints, Glx/tNAA was the most efficient, approaching 80% assurance at approximately 30 participants per group for the 2% threshold (pilot effect+13.6%). tNAA/tCr approached the same level at approximately 50 per group (−8.3%). Gln/tCr showed the largest pilot effect (+24.1%) but did not exceed 80% assurance at any simulated sample size (maximum 76% at N=60). Glu/tCr was the least promising endpoint, with maximum assurance of 39% at N=60 (−3.2%).

## 4. Discussion

This study utilised 7T 3D-CRT-FID-MRSI and sLASER-SVS to investigate neurochemical alterations in ALS across different brain regions, comparing metabolite concentrations between patients and healthy controls, and providing a more detailed metabolic profile than previous studies focused on only single-voxel spectroscopy or 2D-MRSI approaches.^5,24,41^ To ensure regionally comprehensive assessment of metabolite distribution, the analysis was extended beyond sensorimotor ROIs.^59^ The findings highlight widespread neurochemical alterations beyond the traditionally studied motor regions, reinforcing the view that ALS is not limited to motor system degeneration.^22,25,37,41,59,99,100^

### 4.1. Neuronal Integrity and N-Acetylaspartate

A methodological strength enabling these regional findings is the longitudinal segmentation pipeline. By processing all sessions of each participant through a longitudinal template,^88,101^ we maintained anatomical correspondence across visits, and warping multi-modal parcellations preserved fine-grained cortical regional definition that coarser atlases cannot resolve.^89^ Combined with biharmonic spatial smoothing of metabolite maps prior to tCr and tNAA referencing,^92^ this pipeline supports the somatotopic resolution at which our results are reported below.

N-Acetylaspartate is a well-established marker of neuronal integrity, and its reduction in ALS is consistent with neuronal loss and dysfunction.^30^ At baseline, tNAA/tCr was consistently lower in plALS within the sensorimotor strip, with the largest deficits in bilateral precentral and paracentral cortex and extending into postcentral cortex. The motor composite endpoint also showed a clear negative effect This spatial pattern aligns closely with prior MRS studies demonstrating reduced tNAA in motor regions in ALS and is consistent with involvement of cortical motor networks and corticospinal tract degeneration.^22,30,39,100^

The SVS voxel placed in the precentral gyrus may capture progressive neuronal dysfunction in a region tightly linked to disease pathology, whereas the MRSI approach includes a wide set of ROIs where changes may be more heterogeneous and not strictly progressive. Both techniques reinforce the role of reduced tNAA as an indicator of neuronal compromise in ALS, with MRSI adding the advantage of mapping these changes across multiple brain regions simultaneously.

### 4.2. Glutamatergic System and Excitotoxicity in ALS

The findings demonstrate notable elevations in Gln/tCr, Glx/tNAA, and Glu/tCr in multiple motor-related regions of plALS compared to controls. These alterations align with the excitotoxicity hypothesis, which suggests that excessive glutamate levels contribute to neuronal degeneration in ALS.^30,32^ This interpretation is also supported by recent (unpublished) preclinical 7T MRS findings in the SOD1 mouse model of ALS, where increased Gln was reported in motor cortex alongside changes in antioxidant- and energy-related metabolites, including glutathione, taurine, glucose, and tCr.^102^ Though preliminary, these findings support the idea that Gln alterations may reflect broader glutamate-glutamine cycling, antioxidant response, and metabolic stress. Further, our results suggest widespread glutamatergic dysregulation in plALS and align with previous studies reporting elevated Glx in ALS.^5,30,36,43,47,103,104^

One important consideration is that reductions in neuronal volume (as reflected by tNAA reductions) could obscure increases in intracellular Glu concentration, leading to complex metabolic interpretations. Previous research in neurological disorders, including epilepsy, has suggested that using Glu/tNAA as a ratio can partially correct for neuronal volume loss effects, making it a useful measure of glutamatergic abnormalities independent of overall neuronal atrophy.^30,105^ Glx is a composite signal without a single biological meaning and Glu and Gln should be interpreted separately when studying excitotoxicity and neuron-astrocyte cycling.

A central and unanticipated finding of this study is the bulbar-predominant glutamine pattern across the primary motor cortex. All five plALS showed their peak annualised Gln/tCr increase in the bulbar/face zone of the homunculus, irrespective of clinical onset location, including the three lower-limb-onset patients in whom the bulbar zone would be expected to remain relatively preserved on clinical grounds alone. This pattern argues against a simple ‘metabolic mirror’ of clinical onset and instead suggests an intrinsic regional vulnerability of the bulbar motor cortex to glutamatergic dysregulation in ALS, possibly reflecting earlier subclinical involvement of bulbar circuitry, differential baseline glutamatergic metabolism along the homuncular axis, or wider connectivity of the bulbar motor cortex with brainstem and limbic networks. The metabolic-functional coupling within the same somatotopic framework (Glx/tNAA slope vs ALSFRS-R domain decline; r = 0.82) further supports the clinical relevance of this spatial gradient. Confirmatory work in larger cohorts stratified by onset location and disease stage will be required to disentangle these possibilities.

### 4.3. Bayesian assurance and implications for sample size

Less stable or lower-abundance metabolites may require impractically large cohorts to meet stringent decision criteria and are better treated as secondary or exploratory outcomes unless measurement precision is improved. Practically, Glx/tNAA emerges as the most efficient candidate primary endpoint for a confirmatory 7T MRSI cross-sectional study at this disease stage; its metabolite-function coupling (r = 0.82 with matched ALSFRS-R domain decline) and tight posterior uncertainty make it well-suited to a moderate-N trial. tNAA/tCr remains a viable secondary endpoint at moderately larger sample sizes. Gln/tCr, despite the largest pilot effect, requires either modestly larger acquisition voxels (trading some spatial resolution for SNR) or tighter parcel-specific ROI definitions to recover assurance — a measurement-precision design choice rather than a discounting of biological signal, preserving the high-resolution acquisition that enabled the present somatotopic findings.

### 4.4. Limitations

Several additional limitations warrant explicit acknowledgement. First, the cohort was deliberately homogeneous: all five plALS were rapidly progressing (TRICALS −5.8 to - 4.7) and at King’s clinical Stage 2 at first scan, so the present findings may not generalise to slower-progressing or earlier/later-stage patients. Second, all metabolite ratios are referenced to tCr or tNAA rather than absolute concentration; while the tCr-by-zone analysis (3.6) shows the bulbar-predominant Gln/tCr pattern survives denominator drift, absolute quantification remains an important next step. Third, the longitudinal cohort is small and follow-up windows short (3-8 months), which limits the power of group-level interaction tests; this motivates the case-series framing in 3.6 and the assurance-based design guidance in 3.7. Fourth, the somatotopic subdivision of the primary motor cortex used a uniform dorsoventral thirding rather than functional localiser-based boundaries; future work could refine zone boundaries using task-based or tractography-derived homuncular maps.

While session-based comparisons provided insight into metabolic change over time, longitudinal interpretations remain challenging due to disease heterogeneity and the relatively small sample size. One reason for the limited number of plALS was the effort to exclude slow-progressing cases and focus on those in the early stages of the disease, allowing for repeated scans over 3 to 8 months to capture excitotoxicity-related metabolic changes. The small n limited precision and makes ROI-specific estimates sensitive to individual trajectories and prior regularisation. Although hierarchical partial pooling helps stabilise estimation, credible intervals may still understate true uncertainty in settings with sparse participant-level information, and any apparent region-specific effects should be interpreted as exploratory.

Additionally, the lack of a water reference scan for the MRSI sequence limited our ability to perform direct metabolite quantification. Instead, an internal reference (tCr) was used, assuming its stability across plALS and sessions.^106^ Because tCr was used as the reference, changes in creatine could also influence this ratio. While previous literature has suggested creatine is relatively stable in ALS,^106^ our results did not echo these reported trends. This may have been due to inherent variability in the disease cohort, or systematic differences in the acquisition of our disease cohort (see Supplementary materials S2).

From a statistical standpoint, voxelwise models were fitted without explicitly modelling within-ROI spatial autocorrelation, which may inflate the effective sample size and make uncertainty estimates optimistic. The reported motor ‘composite’ was a model-derived pooled summary (averaging ROI-specific posterior draws) rather than a directly modelled participant-level composite endpoint, so it should be interpreted as a pragmatic summary rather than a definitive single-endpoint estimate. Finally, all plALS were taking Riluzole, which can modulate glutamatergic neurotransmission,^37,68,107,108^ and potentially idiosyncratically. Adherence to Riluzole was not measured in this study.

## 5. Conclusion

Using Bayesian mixed-effects models applied to 7T MRSI, we observed consistent group-level differences in markers of neuronal integrity and glutamatergic metabolism across a cortical ROI set, supporting metabolic alterations that extend beyond a single focal motor region in SVS research. Baseline effects were most robust for tNAA/tCr and Gln/tCr, suggesting that glutamine-based measures may provide a more feasible and reproducible glutamatergic endpoint than Glu- or Glx-based ratios in this setting. The ability to separate Glu and Gln at 7T due to its strengths at resolving J-coupled metabolites, strengthens the biological interpretability of these findings. Rather than establishing definitive biomarkers, this study identifies a prioritised set of metabolites and anatomically motivated motor endpoints for future work. These results can inform larger, adequately powered longitudinal studies aimed at confirming region-specific trajectories, clarifying underlying metabolic mechanisms in ALS, and evaluating the clinical utility of MRSI-derived neurochemical markers.

## 6. Data availability

Individual-level neuroimaging data are not publicly deposited because the small cohort size (five plALS, seven non-neurodegenerative controls) precludes meaningful de-identification of MRI. De-identified, voxel-level Glasser-parcellated metabolite ratio data (the inputs to the Bayesian hierarchical models) and the aggregated posterior summaries that underlie every figure and table in this manuscript are available from the corresponding author on reasonable request, subject to a data-sharing agreement consistent with the original ethics approval. Requests should specify the proposed use; access will be granted for non-commercial academic research and will not be granted for re-identification efforts. A redacted summary CSV at the patient-by-zone level (used to produce Supplementary Table S4 and Figure 4C) is included with the publicly released analysis code. A synthetic example dataset that reproduces the data schema (seeded simulation, no real participant information) is included with the analysis code (https://doi.org/10.5281/zenodo.20500484) so the pipeline can be run end-to-end.

## 7. Code availability

All analysis code is openly available under the MIT licence at https://doi.org/10.5281/zenodo.20500484 (archived from https://github.com/thomshaw92/somatotopic-MRSI-MND). The exact version used here is v1.0.0 (https://doi.org/10.5281/zenodo.20500485). The repository contains the full Bayesian modelling pipeline (brms with the cmdstanr backend), figure-generation scripts, and a master rebuild script that regenerates every figure in this manuscript end to end from the de-identified per-voxel CSVs. Software versions are pinned in a renv lockfile and listed in the supplementary methods (R version, brms, cmdstanr, ggsegGlasser, magick, posterior, patchwork, cowplot). Random seeds (seed = 123) are fixed throughout for full numerical reproducibility.

## 8. Acknowledgements

The authors thank all participants who gave their time for this study. The authors thank Kali Chidley, Elaine Kuan, Matilda Gordon, Cory Holdom, Ellen Ritterbusch, and Maja Husum for assistance with administrative tasks during data collection and preparation of this manuscript. We acknowledge Aiman al Najjar, Nicole Atcheson, and the facilities at the Centre for Advanced Imaging, UQ. The authors thank the National Imaging Facility (NIF), Australia. We thank specialist neurology staff at the Royal Brisbane and Women’s staff including Simran Kaur Sarao, Susan Heggie, Lilly Tang, and Rosemary Hanks. We wish to acknowledge The University of Queensland’s Research Computing Centre (RCC) for its support in this research.

## 13 Supplementary Material

Supplementary material is available in a separate file.

