## Supplementary Material for "Glutamine and NAA dissociate in ALS across somatotopically defined motor regions using 7T MRSI"

**Methods S1 - Statistical analysis (full detail)**

S1.1. Software and sampling. All Bayesian models were fitted with brms^96^ using the cmdstanr backend^95,97^. Cross-sectional models (bilateral and lateralised) were sampled with 4 Hamiltonian Monte Carlo chains, 2000 iterations per chain (1000 warmup); longitudinal models with 4 chains, 2500 iterations per chain (1250 warmup). All models used adapt_delta = 0.99 and max_treedepth = 14, with a fixed seed (123) for reproducibility. Within-chain parallelism via threading (8 threads per chain) was used.

S1.2. Prior specifications. For cross-sectional baseline models, weakly informative priors were used: fixed-effect regression coefficients b ~ Normal (0, 0.5); intercept ~ Normal (0, 1); random-effect standard deviations sd ~ Exponential (5); residual scale sigma ~ Exponential (5). For longitudinal models, slightly wider priors were used to accommodate the additional time and interaction terms: b ~ Normal (0, 1); intercept ~ Normal (0, 2); sd ~ Exponential (5); sigma ~ Exponential (5). Priors were centred on no effect with sufficient mass over physiologically plausible ranges to allow the data to dominate; sensitivity to alternative weakly-informative prior specifications was checked informally and did not change the substantive findings.

S1.3. Voxel-cap and IQR filtering. Within each (participant x ROI) cell, retained voxels were sub-sampled with a maximum of 1000 voxels per cell at baseline (cap_voxels with fixed seed) to bound model fitting time without biasing the posterior toward over-represented ROIs. Per-ROI IQR outlier filtering (k = 1.5; pooled across groups within each ROI to avoid removing real group differences) was applied to remove residual artefactual extremes prior to modelling^94^.

S1.4. Bayesian assurance simulation. For each metabolite, 1000 simulated datasets were generated at each of seven candidate per-group sample sizes (N in 10, 15, 20, 25, 30, 40, 60). Each simulation drew a 'true' effect size from the posterior of the pilot motor-composite model and generated voxel-level data under that effect using the pilot residual variance. A simplified single-level brms model (no ROI-varying random effects since simulation operates at the motor-composite level) was refit to each simulated dataset with the same priors as the pilot model. A study was scored as successful if the posterior probability that the implied percent difference exceeded the threshold (2%, 5%, or 10%) was at least 0.95 under a one-sided rule whose direction matched the sign of the pilot effect. Assurance was estimated as the proportion of successful simulations at each N^60,61^.

S1.5. Convergence and model diagnostics. For all fitted models we report Rhat (potential scale reduction factor; target less than 1.01) and bulk and tail effective sample sizes (target greater than 400). Posterior predictive checks used density overlay and grouped summary checks (group SD and upper-quantile checks) to confirm that observed and replicated data were consistent. Leave-one-out cross-validation using Pareto-smoothed importance sampling^109^ was used to identify influential observations (Pareto-k diagnostics; observations with k greater than 0.7 flagged for inspection). All models passed convergence checks (Rhat less than 1.01, ESS greater than 400) except for one parameter in the bilateral Glx/tNAA baseline model (Rhat = 1.011, marginal); refitting with longer warmup did not change the substantive findings.

S1.6. Software versions and reproducibility. Analyses were run in R (R version 4.5.3 (2026-03-11)) using brms^96^, cmdstanr, posterior, patchwork, cowplot, ggseg2 / ggsegGlasser^110^. Full analysis code and the seed (123) used for reproducibility are available at <https://doi.org/10.5281/zenodo.20500484> (archived from <https://github.com/thomshaw92/somatotopic-MRSI-MND> ).

S1.7. Methodological notes. Our SVS/MRSI scans could be biased due to subject motion and other instabilities. Previous studies have demonstrated that motion correction improves data quality in high-resolution 3D-CRT-based FID-MRSI at 3T^111^, and its integration at 7T, particularly given the longer 15-minute scan duration because of higher spatial resolution, could enhance spectral reliability and spatial precision. Future studies should evaluate the feasibility of incorporating real-time motion correction at 7T, as the longer acquisition times make MRSI particularly sensitive to motion. The real-time correction framework currently implemented at 3T is not yet validated for 7T CRT-FID-MRSI, but its development would likely improve spectral reliability and may help narrow the reproducibility gap between 3 T and 7T.

**Supplementary Results S2**

**S2.1. MRSI**

For MRSI spectral quality at the voxel level was evaluated by calculating the SNR of the NAA peak using the pseudo-replica method, along with measuring the full width at half maximum (FWHM) of total creatine (tCr: Cr+PCr) peak at 3.02 ppm^74^ and for spectral quality comparisons between the ALS and NC groups, the SNR and FWHM from all voxels within each ROI were averaged to obtain the mean SNR and mean FWHM per ROI. For SVS spectra quality, SNR and FWHM of tCr were extracted from Osprey.

High resolution and quality metabolite maps and spectra were obtained at 7T using 3D-CRT-FID-MRSI from plALS and NCs (Supplementary Figure S5). The quality of MRSI spectra and fitting is shown in Supplementary Figure S5. ROI-based comparisons showed that the mean SNR was lower in plALS in 22 out of 26 ROIs, while FWHM was higher in 20 out of 26 ROIs compared to NCs. Significant differences (p < 0.05) were observed in six ROIs, including the superior frontal, precentral, and paracentral regions in both hemispheres. When considering the entire MRSI FOV, the mean SNR across all ROIs was 16.80 ± 4.09 in plALS and 18.67 ± 4.06 in NCs, while the mean FWHM was 0.064 ± 0.020 ppm in plALS and 0.058 ± 0.012 ppm in NCs.

**S2.2. SVS**

Supplementary Figure S6 shows representative spectra from a pALS and a NC obtained from the precentral gyrus along with LCModel fits and residuals. For this voxel SNR as assessed for tCr was 157.6 ± 26.1 for plALS and 165.6 ± 18.8 for NC (difference was not significant). The linewidth (tCr FWHM) was 0.039 ± 0.002 ppm for plALS and 0.04 ± 0.004 ppm for NC (not significant).

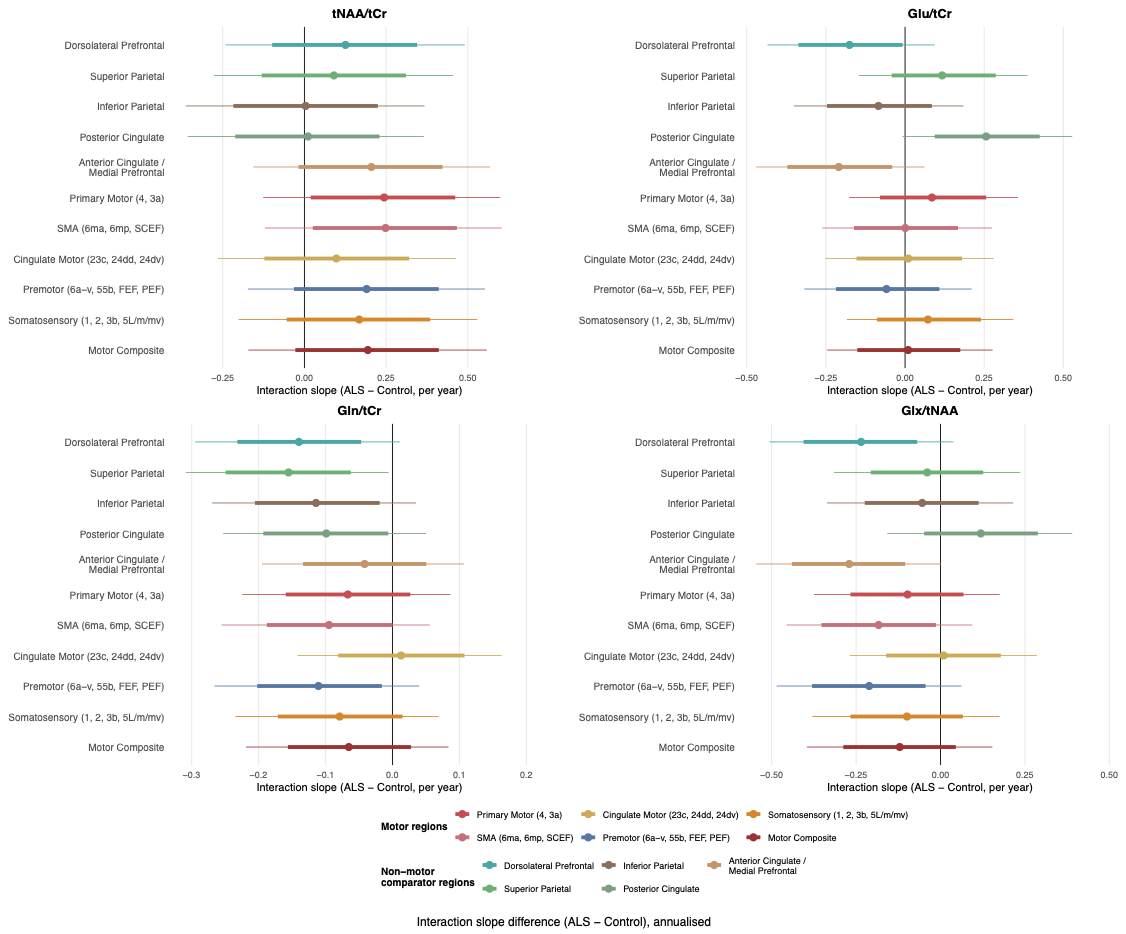

**Supplementary Figure S1: Longitudinal interaction-slope forest plots for the four metabolite ratios.** Posterior median and 95% credible interval for the ROI-specific Group x Time interaction (annualised slope difference, plALS minus NCs) for tNAA/tCr, Glu/tCr, Gln/tCr, and Glx/tNAA.

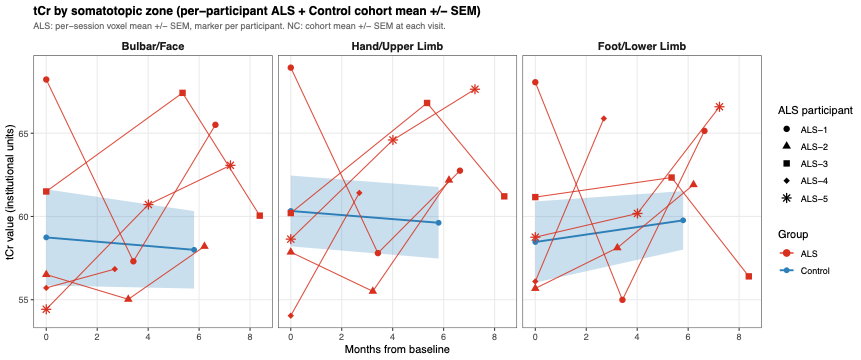

**Supplementary Figure S2. tCr by somatotopic zone (ALS + non-neurodegenerative controls).** Per-session tCr value (institutional units) split into bulbar/face, hand/upper-limb, and foot/lower-limb zones of the bilateral primary motor cortex (Glasser parcels 4 and 3a). plALS are plotted individually as per-participant trajectories using distinct point markers (ALS-1 to ALS-5; matching Table 2), with per-session error bars representing the within-participant standard error of the mean across voxels in that zone. Non-neurodegenerative controls (NCs) are aggregated into a cohort trajectory: mean ± SEM across NC participants at each session, shown as a line with a shaded ribbon. NC visit means are plotted at the cohort-average for that visit; visits with fewer than two contributing NCs are omitted.

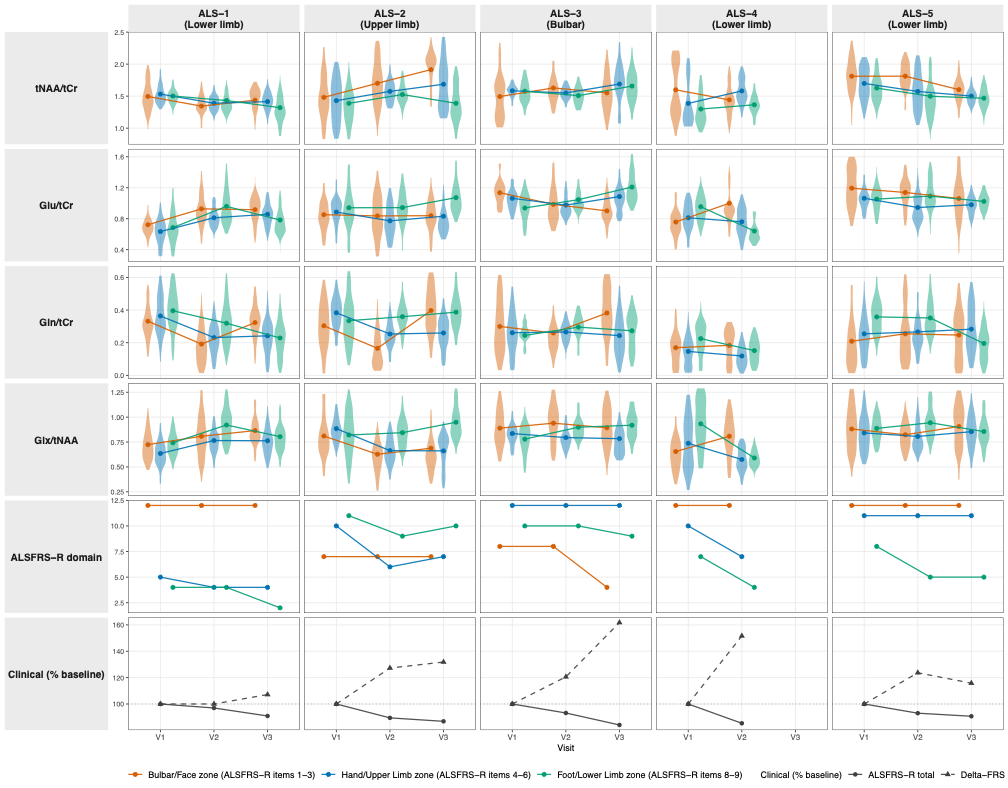

**Supplementary Figure S3. Per-participant longitudinal somatotopic profiles.** For each plALS (ALS-1 to ALS-5; columns), each row shows: tNAA/tCr, Glu/tCr, Gln/tCr, and Glx/tNAA voxel-level distributions across the bilateral primary motor cortex (Glasser parcels 4 + 3a) split into bulbar/face, hand/upper-limb, and foot/lower-limb tertiles along the dorsoventral axis; ALSFRS-R domain subscores (Bulbar items 1–3, Fine Motor 4–6, Gross Motor 8–9) coloured by matched somatotopic zone; and global clinical metrics expressed as percent of session-1 baseline (ALSFRS-R total and Delta-FRS). Violins show per-voxel value distributions per session per zone after Tukey IQR (k=1.5) outlier filtering and a cap of 150 voxels per cell. Dotted line in the clinical row marks 100% (no change from baseline). Onset location is given beneath each participant identifier.

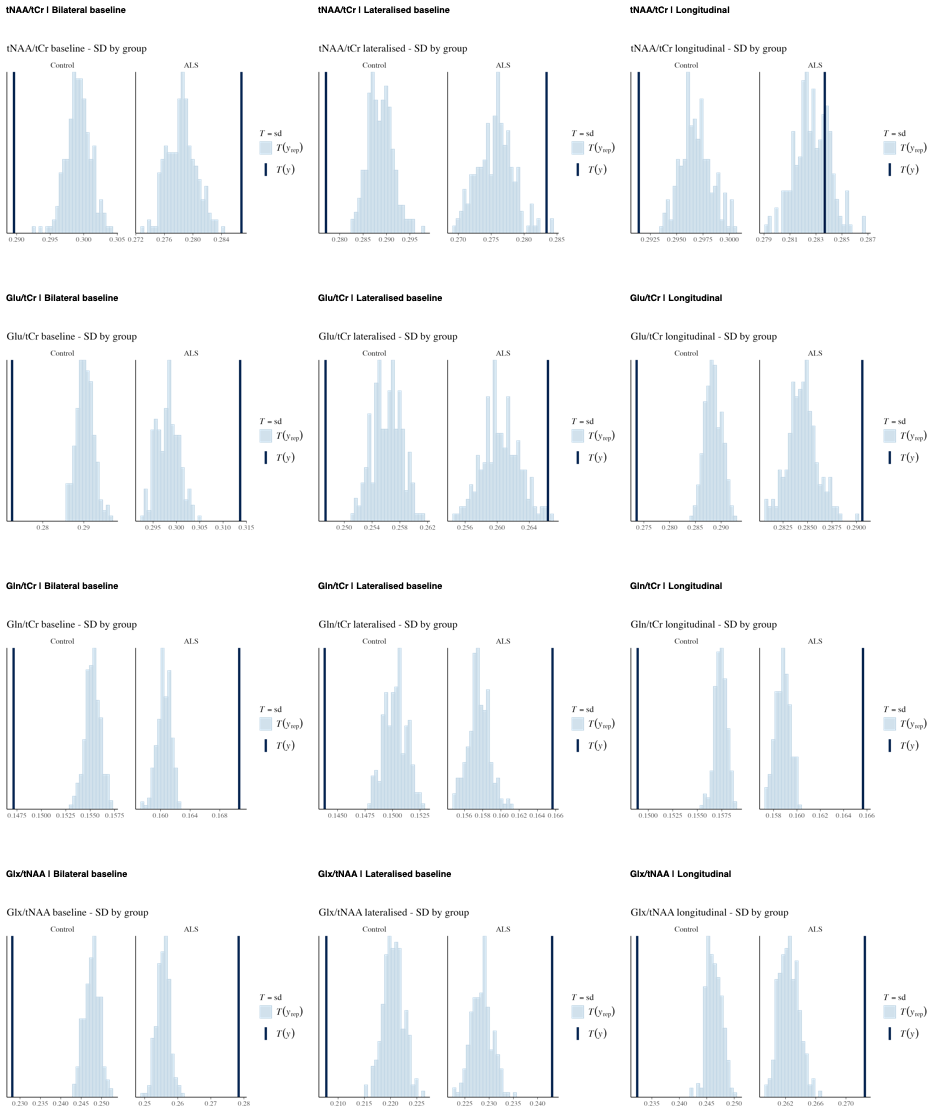
**Supplementary Figure S4. Posterior predictive checks for all twelve fitted Bayesian hierarchical models.** Each panel shows the group-stratified standard-deviation check from brms::pp_check(type = 'stat_grouped', stat = 'sd', group = 'Group'): bars give the SD of replicated voxel-level draws for plALS and NCs separately, and the vertical line marks the SD of the observed data. Panels are arranged with metabolite ratio in rows (tNAA/tCr, Glu/tCr, Gln/tCr, Glx/tNAA) and model type in columns (bilateral baseline, lateralised baseline, longitudinal Group x Time).

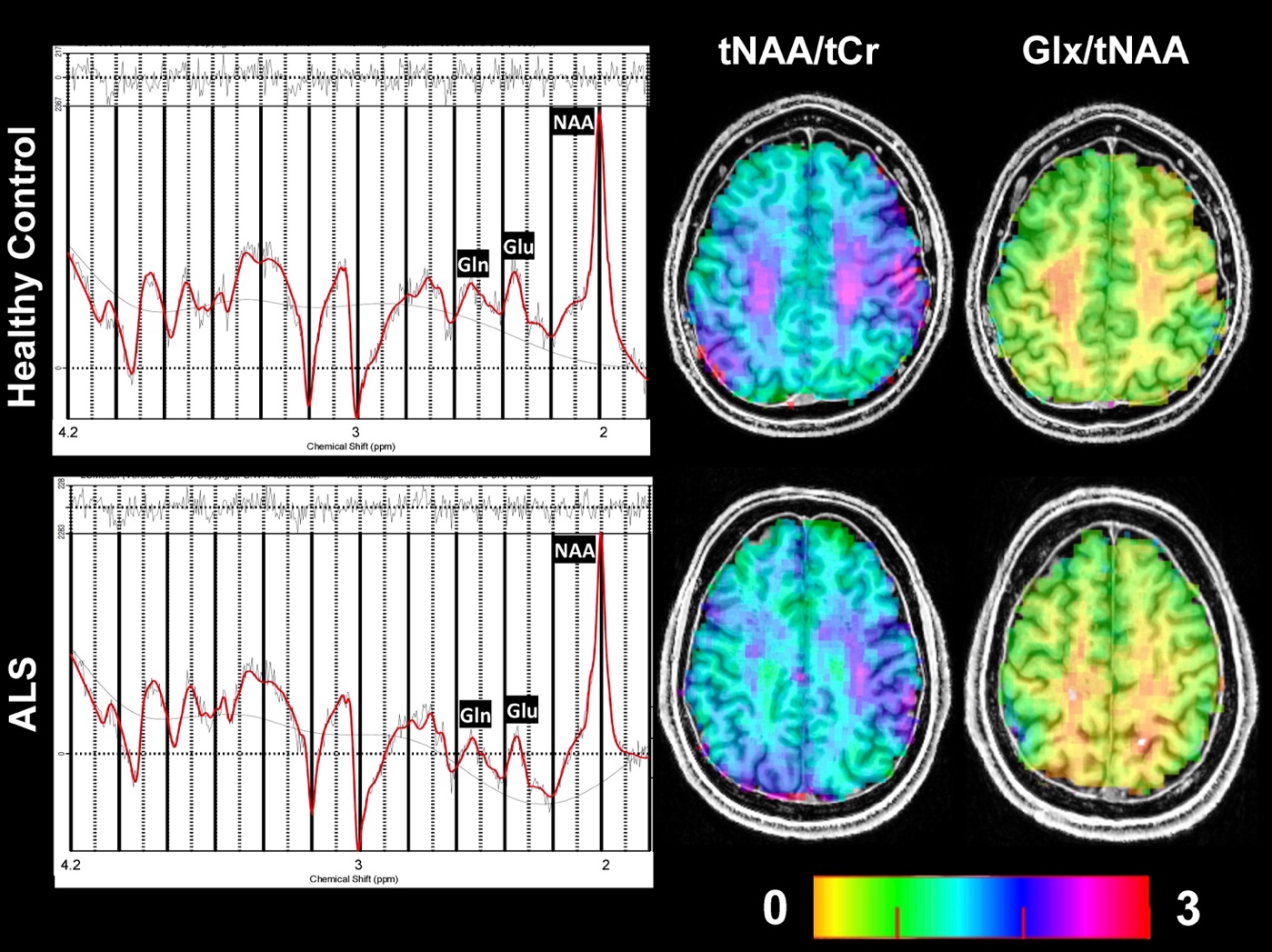

Supplementary Figure S5. Representative 1H MR Spectra and LCModel fitting results for representative data from a NC and one plALS measured at 7T using FID-CRT-MRSI.

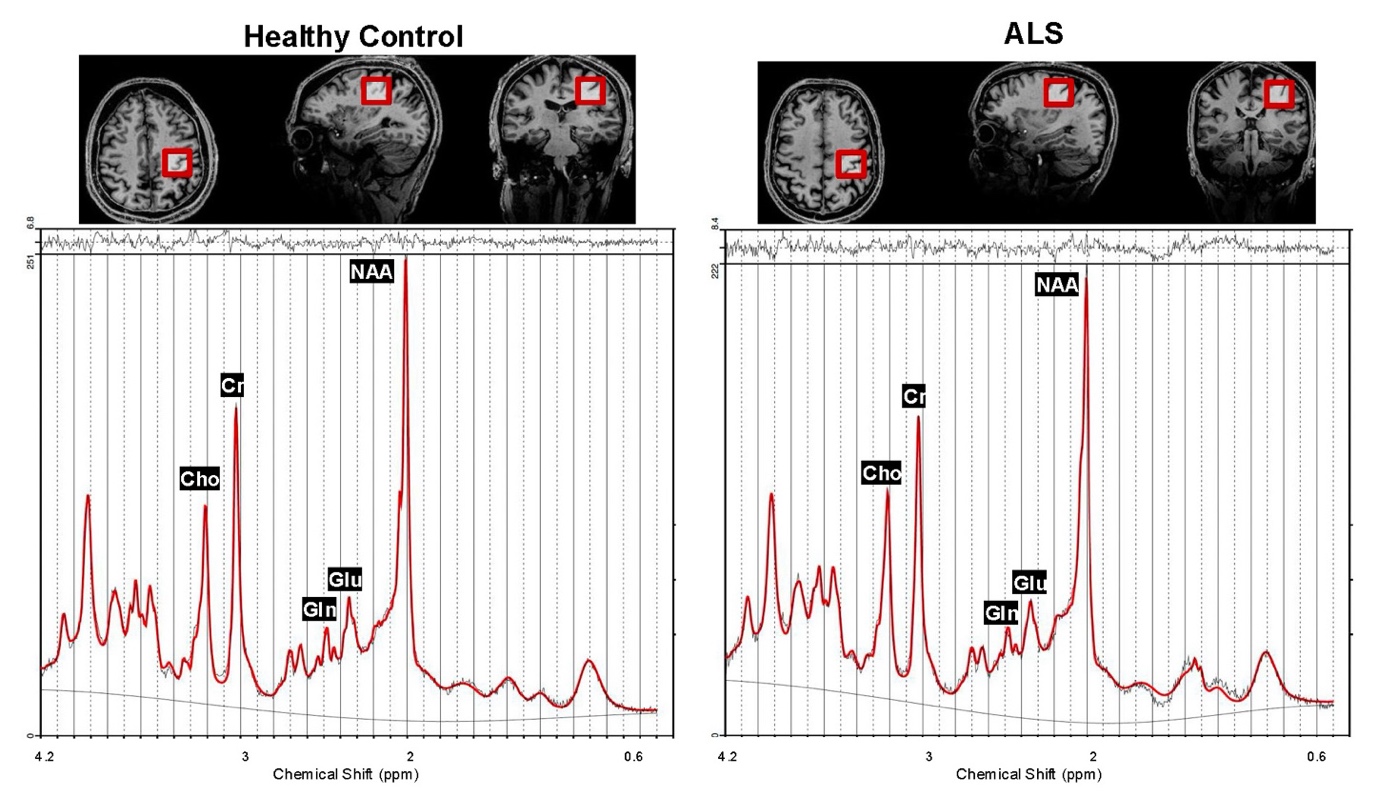

Supplementary Figure S6: Representative 1H MR Spectra and LCModel fitting results for representative data from a NC and one plALS measured at 7T from the precentral gyrus with the sLASER sequence (TE=26 ms, TR=8 s).

Table S1: The MRSinMRS Checklist^81^. A standardised reporting framework for in vivo magnetic resonance spectroscopy studies, completed for the present 7T 3D-CRT-FID-MRSI and sLASER-SVS acquisitions. Each row reports on a recommended methodological item covering hardware, sequence parameters, acquisition, post-processing, and quantification decisions described in Methods 2.2 and 2.

| 1. Hardware | | |
| --- | --- | --- |
| a. Field strength (T) | 7 | |
| b. Manufacturer | Siemens | |
| c. Model | vd syngo MR E12 | |
| d. RF coils | 32 ch ^1^H head coil | |
| 2. Acquisition | | |
| a. Pulse sequence | CRT-FID-MRSI | sLASER |
| b. VOI locations | Cerebrum | Right Precentral Gyrus |
| c. Voxel size (mm^3^) | 3.4×3.4×3.5 | 25×25×25 |
| d. TR/TE (ms) | 460/1.3 | 8000/26 |
| e. Averages | 1 | 16 |
| f. spectral width in Hz, number of spectral points; If MRSI: 2D or 3D, FOV in all directions, matrix size, acceleration factors, sampling method | BW 2778, MRSI: 3D, 220×220×110 mm3 , 64×64×31, spatial-spectral encoding | BW 6000, 2048 spectral points |
| g. Water suppression method | WET | VAPOR |
| h. Shimming method | Standard shim + manual adjustment, water peak < 50 Hz | fastestmap |
| 3. Data analysis methods and outputs | | |
| a. Analysis software | LCModel 6.3-1 | Osprey |
| b. Processing steps deviating from quoted reference or product | Internal water reference | Default Osprey |
| c. Output measure | Ratio/tCr | |
| d. Quantification references and assumptions, fitting model assumptions | Simulated in NMRScope-B, macromolecular background | Basis set includes 19 simulated metabolites + measured macromolecule. LCModel basline knot spacing 5.00 ppm |
| 4. Data quality | | |
| a. Reported variables (SNR, linewidth (with reference peaks)) | MRSI: SNR for NAA was estimated voxel-wise using the pseudo-replica method, which incorporates receiver noise prescans acquired at the start of the MRSI sequence. The FWHM of the spectral peaks for tCr at 3.02 ppm was calculated from the LCModel fits. sLASER: SNR dividing the tCr by the standard deviation of noise within the range of -2 to 0 ppm. Linewidth FWHM of a Lorentzian peak model for the water peak between 4.4 and 5.0 ppm | |
| b. Data exclusion criteria | CRLB > 50% | |
| c. Quality measures of postprocessing model fitting (eg CRLB, goodness of fit, SD of residual) | CRLB, and SD of residual | |
| d. Sample spectrum | (Supplementary Figure S6). | |

Supplementary Table S2: Longitudinal fixed-effect estimates for the four metabolite ratios. Posterior median and 95% credible interval for the population-average Group, Time, and Group x Time interaction terms from the longitudinal hierarchical models.

| Metabolite | Group (ALS vs Control) | Timepoint | Group x Time Interaction |
| --- | --- | --- | --- |
| tNAA/tCr | -0.07267 (-0.21373, 0.06786) | -0.00354 (-0.02491, 0.01833) | 0.01153 (-0.01910, 0.04247) |
| Glu/tCr | -0.03153 (-0.17734, 0.11073) | 0.00335 (-0.01383, 0.01911) | 0.00024 (-0.02306, 0.02426) |
| Gln/tCr | 0.03743 (-0.03808, 0.11042) | 0.00292 (-0.00606, 0.01222) | -0.00736 (-0.02069, 0.00530) |
| Glx/tNAA | 0.05227 (-0.07489, 0.18337) | 0.00544 (-0.01073, 0.02070) | -0.00889 (-0.03271, 0.01542) |

Supplementary Table S3: Per-patient longitudinal motor-composite metabolite trajectories. Absolute and annualised changes in tNAA/tCr, Glu/tCr, Gln/tCr, Glx/tNAA, and tCr over each patient's available follow-up window, derived from the per-patient case-series analysis. Provides the per-patient detail underlying the cohort summaries in 3.6. Motor composite = bilateral Primary Motor + Premotor + SMA + Cingulate Motor. 'Stable' is defined as |% change over interval| < 1% AND |annualised slope| < 0.05/yr.

| **Participant** | **Clinical onset** | **Baseline ALSFRS-R total** | **Delta-FRS** | **Visits (N)** | **Follow-up (months)** | **Metabolite** | **% change over interval** | **Annualised slope (per year)** | **Direction** |
| --- | --- | --- | --- | --- | --- | --- | --- | --- | --- |
| ALS-1 | Lower limb | 33 | 0.28 | 3 | 7 | tNAA/tCr | -3.5 | -0.093 | Declined |
| ALS-1 | Lower limb | 33 | 0.28 | 3 | 7 | Glu/tCr | +22.0 | +0.334 | Increased |
| ALS-1 | Lower limb | 33 | 0.28 | 3 | 7 | Gln/tCr | -17.3 | -0.129 | Declined |
| ALS-1 | Lower limb | 33 | 0.28 | 3 | 7 | Glx/tNAA | +13.6 | +0.207 | Increased |
| ALS-1 | Lower limb | 33 | 0.28 | 3 | 7 | tCr | -5.8 | -7.023 | Declined |
| ALS-2 | Upper limb | 38 | 0.22 | 3 | 6 | tNAA/tCr | +11.6 | +0.341 | Increased |
| ALS-2 | Upper limb | 38 | 0.22 | 3 | 6 | Glu/tCr | -4.3 | -0.084 | Declined |
| ALS-2 | Upper limb | 38 | 0.22 | 3 | 6 | Gln/tCr | +11.9 | +0.066 | Increased |
| ALS-2 | Upper limb | 38 | 0.22 | 3 | 6 | Glx/tNAA | -9.2 | -0.154 | Declined |
| ALS-2 | Upper limb | 38 | 0.22 | 3 | 6 | tCr | +4.5 | +4.849 | Increased |
| ALS-3 | Bulbar | 44 | 0.34 | 3 | 8 | tNAA/tCr | -3.1 | -0.070 | Declined |
| ALS-3 | Bulbar | 44 | 0.34 | 3 | 8 | Glu/tCr | -5.2 | -0.082 | Declined |
| ALS-3 | Bulbar | 44 | 0.34 | 3 | 8 | Gln/tCr | +20.5 | +0.091 | Increased |
| ALS-3 | Bulbar | 44 | 0.34 | 3 | 8 | Glx/tNAA | +6.6 | +0.086 | Increased |
| ALS-3 | Bulbar | 44 | 0.34 | 3 | 8 | tCr | -5.5 | -4.968 | Declined |
| ALS-4 | Lower limb | 41 | 0.62 | 2 | 3 | tNAA/tCr | +3.8 | +0.245 | Increased |
| ALS-4 | Lower limb | 41 | 0.62 | 2 | 3 | Glu/tCr | +0.7 | +0.025 | Stable |
| ALS-4 | Lower limb | 41 | 0.62 | 2 | 3 | Gln/tCr | -17.2 | -0.177 | Declined |
| ALS-4 | Lower limb | 41 | 0.62 | 2 | 3 | Glx/tNAA | -9.5 | -0.321 | Declined |
| ALS-4 | Lower limb | 41 | 0.62 | 2 | 3 | tCr | +5.2 | +13.325 | Increased |
| ALS-5 | Lower limb | 43 | 0.38 | 3 | 7 | tNAA/tCr | -5.7 | -0.156 | Declined |
| ALS-5 | Lower limb | 43 | 0.38 | 3 | 7 | Glu/tCr | -6.9 | -0.135 | Declined |
| ALS-5 | Lower limb | 43 | 0.38 | 3 | 7 | Gln/tCr | -10.0 | -0.056 | Declined |
| ALS-5 | Lower limb | 43 | 0.38 | 3 | 7 | Glx/tNAA | -3.1 | -0.050 | Declined |
| ALS-5 | Lower limb | 43 | 0.38 | 3 | 7 | tCr | +14.5 | +13.639 | Increased |

Supplementary Table S4: Sensitivity analysis - participant-mean metabolite-function correlations. The headline metabolite-function correlations reported in Section 3.6 treat each of the 15 patient-zone pairs as independent. As a sensitivity analysis, the same correlations were recomputed after collapsing each patient's three zones to a participant-mean slope (n=5 participants). Convergent direction and rank-ordering between the n=15 and n=5 analyses support robustness; divergence indicates clustering effects. Glx/tNAA (convergent) shaded.

| **Metabolite** | **n** | **r** | **p_value** | **level** |
| --- | --- | --- | --- | --- |
| Gln/tCr | 5 | 0.34 | 0.575 | participant_mean (n=5) |
| Gln/tCr | 15 | 0.597 | 0.0188 | patient_zone (n=15) |
| Glu/tCr | 5 | 0.553 | 0.334 | participant_mean (n=5) |
| Glu/tCr | 15 | 0.67 | 0.00632 | patient_zone (n=15) |
| Glx/tNAA | 5 | 0.927 | 0.0234 | participant_mean (n=5) |
| Glx/tNAA | 15 | 0.822 | 1.68E-04 | patient_zone (n=15) |
| tNAA/tCr | 5 | -0.332 | 0.585 | participant_mean (n=5) |
| tNAA/tCr | 15 | -0.511 | 0.0517 | patient_zone (n=15) |

*Supplementary Table S5:* Baseline ROI-wise group differences (plALS minus NCs, expressed as percent of NC mean) from Bayesian hierarchical mixed-effects models with Student-t likelihood. P denotes the posterior probability of group separation in the indicated direction.

| **Metabolite** | **ROI** | **Region type** | **Effect (median, 95% CrI)** | **P** |
| --- | --- | --- | --- | --- |
| *NAA/tCr* | Somatosensory (1, 2, 3b, 5L/m/mv) | Non-motor | -7.8% (-15.2 to +0.2) | P (<0) = 0.97 |
|  | Superior Parietal | Non-motor | -3.7% (-11.8 to +4.9) | P (<0) = 0.83 |
|  | Dorsolateral Prefrontal | Non-motor | -5.1% (-13.1 to +3.1) | P (<0) = 0.90 |
|  | Anterior Cingulate / Medial Prefrontal | Non-motor | -6.0% (-14.4 to +2.9) | P (<0) = 0.92 |
|  | Inferior Parietal | Non-motor | -4.3% (-12.4 to +4.2) | P (<0) = 0.85 |
|  | Posterior Cingulate | Non-motor | -0.0% (-9.2 to +9.8) | P (<0) = 0.50 |
| *Glu/tCr* | Somatosensory (1, 2, 3b, 5L/m/mv) | Non-motor | -3.4% (-17.3 to +12.8) | P (<0) = 0.70 |
|  | Superior Parietal | Non-motor | -0.3% (-14.6 to +16.7) | P (<0) = 0.52 |
|  | Dorsolateral Prefrontal | Non-motor | +1.9% (-11.4 to +18.0) | P (<0) = 0.38 |
|  | Anterior Cingulate / Medial Prefrontal | Non-motor | -0.0% (-13.8 to +16.2) | P (<0) = 0.50 |
|  | Inferior Parietal | Non-motor | +1.7% (-11.8 to +17.8) | P (<0) = 0.39 |
|  | Posterior Cingulate | Non-motor | -2.6% (-18.1 to +15.8) | P (<0) = 0.63 |
| *Gln/tCr* | Somatosensory (1, 2, 3b, 5L/m/mv) | Non-motor | +24.4% (-7.4 to +64.7) | P (>0) = 0.94 |
|  | Superior Parietal | Non-motor | +33.2% (-0.7 to +75.7) | P (>0) = 0.97 |
|  | Dorsolateral Prefrontal | Non-motor | +37.5% (+10.7 to +69.9) | P (>0) = 1.00 |
|  | Anterior Cingulate / Medial Prefrontal | Non-motor | +11.0% (-11.1 to +36.2) | P (>0) = 0.84 |
|  | Inferior Parietal | Non-motor | +37.0% (+1.4 to +84.4) | P (>0) = 0.98 |
|  | Posterior Cingulate | Non-motor | +22.8% (-9.8 to +64.9) | P (>0) = 0.92 |
| *Glx/tNAA* | Somatosensory (1, 2, 3b, 5L/m/mv) | Non-motor | +11.2% (-1.5 to +25.8) | P (>0) = 0.96 |
|  | Superior Parietal | Non-motor | +10.5% (-2.1 to +25.3) | P (>0) = 0.95 |
|  | Dorsolateral Prefrontal | Non-motor | +13.8% (+2.4 to +26.7) | P (>0) = 0.99 |
|  | Anterior Cingulate / Medial Prefrontal | Non-motor | +9.7% (-0.8 to +21.8) | P (>0) = 0.96 |
|  | Inferior Parietal | Non-motor | +10.6% (-0.4 to +23.2) | P (>0) = 0.97 |
|  | Posterior Cingulate | Non-motor | +5.4% (-6.8 to +19.2) | P (>0) = 0.83 |
